# Prospective SARS-CoV-2 Booster Vaccination in Immunosuppressant-Treated Systemic Autoimmune Disease Patients in a Randomized Controlled Trial

**DOI:** 10.1101/2025.03.25.25324558

**Authors:** Meggan Mackay, Catriona A Wagner, Ashley Pinckney, Jeffrey A Cohen, Zachary S Wallace, Arezou Khosroshahi, Jeffrey A Sparks, Sandra Lord, Amit Saxena, Roberto Caricchio, Alfred HJ Kim, Diane L Kamen, Fotios Koumpouras, Anca D Askanase, Kenneth Smith, Joel M Guthridge, Gabriel Pardo, Yang Mao-Draayer, Susan Macwana, Sean McCarthy, Matthew A Sherman, Sanaz Daneshfar Hamrah, Maria Veri, Sarah Walker, Kate York, Sara Tedeschi, Jennifer Wang, Gabrielle Dziubla, Mike Castro, Robin Carroll, Sandeep Narpala, Bob C Lin, Leonid Serebryannyy, Adrian McDermott, ACV01 Study Team, William T Barry, Ellen Goldmuntz, James McNamara, Aimee S. Payne, Amit Bar-Or, Dinesh Khanna, Judith A James

## Abstract

**Background.:** Autoimmune disease patients on immunosuppressants exhibit reduced humoral responses to primary COVID-19 vaccination. Booster vaccine responses and the effects of holding immunosuppression around vaccination are less studied. We evaluated the efficacy and safety of additional vaccination in mycophenolate mofetil/mycophenolic acid (MMF/MPA)-, methotrexate (MTX)-, and B cell-depleting therapy (BCDT)-treated autoimmune disease patients, including the impact of withholding MMF/MPA and MTX.

**Methods.:** In this open-label, multicenter, randomized trial, 22 MMF/MPA-, 26 MTX-, and 93 BCDT-treated autoimmune disease patients with negative or suboptimal antibody responses to initial COVID-19 vaccines (BNT162b2, mRNA-1273, or AD26.COV2.S) received a homologous booster. MMF/MPA and MTX participants were randomized (1:1) to continue or withhold treatment around vaccination. The primary outcome was the change in anti-Wuhan-Hu-1 receptor-binding domain (RBD) concentrations at 4 weeks post-additional vaccination. Secondary outcomes included adverse events, COVID-19 infections, and autoimmune disease activity through 48 weeks.

**Results.:** Additional vaccination increased anti-RBD concentrations in MMF/MPA and MTX patients, irrespective of whether immunosuppression was continued or withheld. BCDT-treated patients also demonstrated increased anti-RBD concentrations, albeit lower than MMF/MPA- and MTX-treated cohorts. COVID-19 infections occurred in 30-46% of participants, were predominantly mild, and included only two non-fatal hospitalizations. Additional vaccination was well-tolerated, with low frequencies of severe disease flares and adverse events.

**Conclusion.:** Additional COVID-19 vaccination is effective and safe in immunosuppressant-treated autoimmune disease patients, regardless of whether MMF/MPA or MTX is withheld.

**Trial Registration.** ClinicalTrials.gov (NCT#05000216)

## Introduction

Autoimmune disease patients treated with immunosuppressants (IS), such as methotrexate (MTX), mycophenolate mofetil/mycophenolic acid (MMF/MPA), and B cell-depleting therapies (BCDT), exhibit impaired humoral responses following initial COVID-19 vaccination series, which is associated with an increased risk of breakthrough infections (1–3). Therefore, improving vaccine efficacy in individuals with an inadequate humoral immune response is crucial. In BCDT- and MMF/MPA-treated autoimmune disease patients, booster vaccination induces a humoral response in some seronegative patients, but seropositivity remains at a lower frequency and lower levels compared to controls (4–7). In MTX-treated autoimmune disease patients, seroconversion is relatively high following the primary vaccine series; however, antibody levels are decreased compared to controls (8). While an additional vaccination further increases anti-SARS-CoV-2 antibody levels, conflicting data exists on whether antibody levels remain lower compared to controls (5,7,9–12). Furthermore, prospective clinical trial data on the effects of additional vaccination on breakthrough infections in MTX-, MMF/MPA-, and BCDT-treated patients and clinical trials addressing the immunogenicity of booster vaccination in these populations are lacking.

MMF/MPA and MTX can impair vaccine responses, and current guidelines recommend withholding MMF/MPA and MTX for 1-2 weeks around each COVID-19 vaccine dose (13). However, there are limited data from clinical trials demonstrating the benefit and safety of this approach following booster vaccination. In the VROOM clinical trial, a 2-week hold of MTX following additional vaccination with BNT162b2 (Pfizer–BioNTech), mRNA-1273 (Moderna), or AZD1222 (AstraZeneca) vaccines in patients with immune-mediated inflammatory disease resulted in a 2.19-fold increase in antibodies against the receptor-binding domain (RBD) compared to patients who continued MTX (14). This response was sustained through 26 weeks post-booster vaccination (15). Although withholding MTX temporarily increased self-reported disease flares, the majority of flares were limited, self-managed, and did not require medical attention, with no serious adverse events (AEs) or deaths (14,15). Despite improved humoral responses, new COVID-19 infections throughout the VROOM trial occurred at similar frequencies and severity regardless of MTX suspension (15). While less is known about the effects of MMF/MPA withdrawal on SARS-CoV-2 vaccine responses in autoimmune disease patients, an observational study of 195 rheumatic and musculoskeletal disease patients demonstrated that MMF/MPA withholding near the time of vaccination increased the frequency and titer of anti-RBD antibodies (16). Indeed, limited clinical trial data supports current recommendations regarding immunosuppressive treatment withhold, especially regarding clinical vaccine efficacy and the effects in patients with inadequate or modest humoral responses to the primary vaccine series.

In this randomized, multi-site, open-label clinical trial, we determined the immunogenicity, safety, and clinical efficacy of a homologous additional COVID-19 vaccine dose in autoimmune disease patients on MTX, MMF/MPA, or BCDT with suboptimal responses to the primary COVID-19 vaccine series. In addition, we evaluated the impacts of temporarily withholding MTX or MMF/MPA on the vaccine response.

## Results

### Study participants

Between August 2021 and August 2022, we screened 279 participants for Stage 1, of whom 148 were randomized or allocated; this included 25 MMF/MPA- (11 continue, 14 withhold), 28 MTX- (14 continue, 14 withhold), and 95 BCDT-treated (Figure 1). Of these participants, 22 MMF/MPA-treated (10 continue, 12 withhold), 26 MTX-treated (13 continue, 13 withhold), and 93 BCDT-treated participants received an additional vaccine. In total, 82 BNT162b2, 48 mRNA-1273, and 10 AD26.COV2.S vaccines were received. Due to the small sample sizes, participants who received either BNT162b2 (Pfizer–BioNTech) or mRNA-1273 (Moderna) were analyzed together, while those who received the AD26.COV2.S (Janssen) vaccine were analyzed separately.

**Figure 1.**
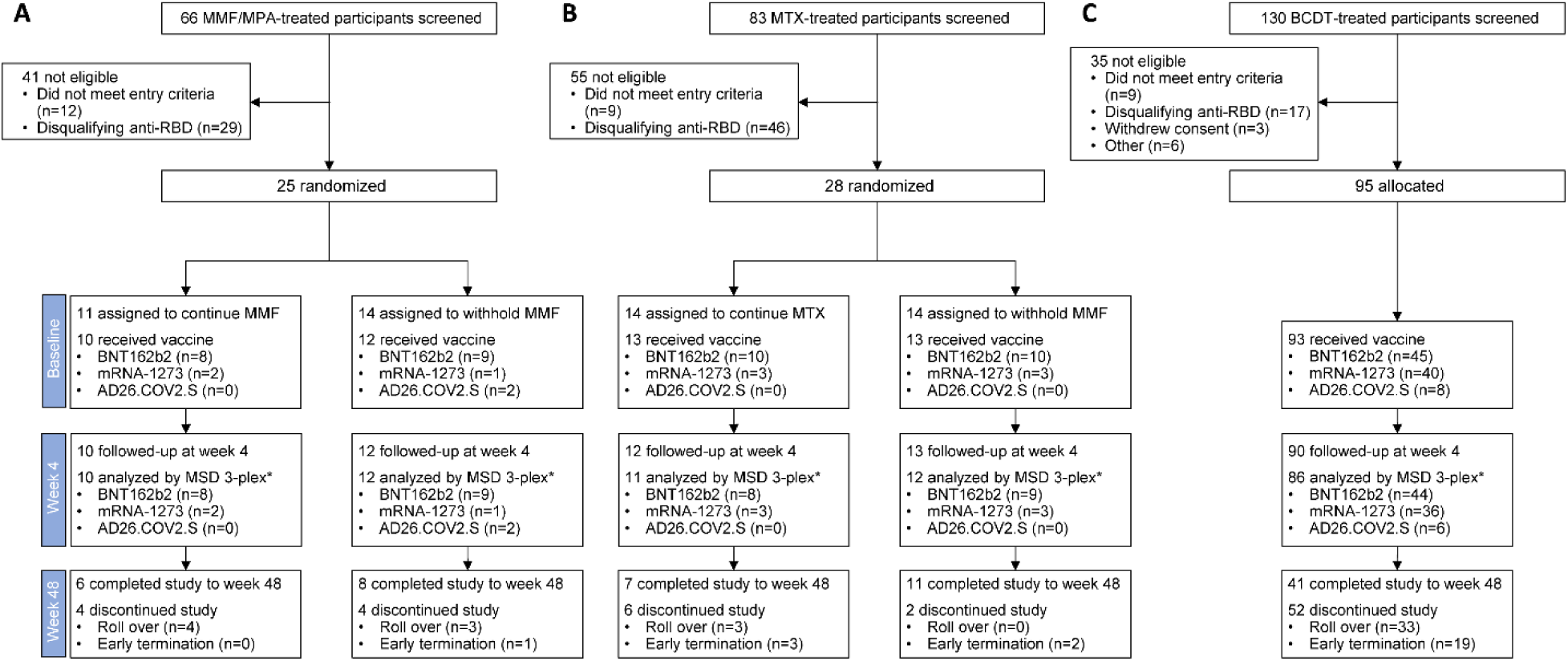
ACV01 trial profile. MMF, mycophenolate mofetil; MPA, mycophenolic acid; MTX, methotrexate; BCDT, B cell-depleting therapy. *Samples were excluded from the humoral response analyses if the participant experienced a COVID-19 infection, monoclonal antibody use, or a COVID-19 vaccination off-study between the baseline and week 4 visit. **Some individuals were rolled over to Stage 2 of this study to assess booster with a non-homologous vaccine.

Baseline demographics were similar between cohorts in the vaccinated population. Most participants were women (MMF/MPA, 91%; MTX, 96%; BCDT, 71%), with the majority identifying as White or Black and non-Hispanic or Latino (Table 1). The median age for all cohorts was 54 years. The MMF/MPA cohort consisted of 73% (n=16) systemic lupus erythematosus (SLE), 23% (n=5) systemic sclerosis (SSc), and 5% (n=1) pemphigus subjects, while the MTX cohort consisted of 77% (n=20) rheumatoid arthritis (RA) and 23% (n=6) SLE (Table 1). Most participants in the BCDT cohort had multiple sclerosis (MS) (72%, n=67) and RA (19%, n=18). Only 1 (5%) MMF/MPA-treated and 7 (8%) BCDT-treated participants had a prior COVID-19 infection as self-reported at screening. Few participants in the MMF/MPA- (n=9; 41%) and MTX-treated (n=1; 4%) cohorts exhibited a negative humoral response to the initial COVID-19 vaccination, while the majority in the BCDT cohort (n=69; 74%) had a negative humoral response (Table 1). Most participants taking MMF/MPA had SLE with mean doses of 2375±806.2 mg daily for MMF and 900±763.7 mg daily for MPA. Most participants taking MTX had RA with a mean dose of 17±5.6 mg weekly (Supplemental Table 1). The majority of participants with MS were treated with ocrelizumab (n=62), and a few were on ofatumumab (n=3). Most of the participants receiving rituximab had RA (n=18). The mean steroid dose was 4-6 mg daily across the participants receiving prednisone (SLE, n=14; RA, n=6; pemphigus, n=1) (Supplemental Table 1).

**Table 1.** Baseline demographics and characteristics in the vaccinated population.

|  | MMF/MPA<br>(n=22) | MTX<br>(n=26) | BCDT<br>(n=93) |
| --- | --- | --- | --- |
| **Sex, n (%)** |  |  |  |
| Female | 20 (91) | 25 (96) | 66 (71) |
| Male | 2 (9) | 1 (4) | 27 (29) |
| **Race, n (%)** |  |  |  |
| White | 11 (50) | 16 (62) | 78 (83) |
| Black | 10 (46) | 5 (19) | 12 (13) |
| Asian | 0 | 2 (8) | 1 (1) |
| American Indian or Alaska Native | 0 | 1 (4) | 0 |
| Multiple | 0 | 1 (4) | 1 (1) |
| Unknown or not reported | 1 (5) | 1 (4) | 1 (1) |
| **Ethnicity, n (%)** |  |  |  |
| Hispanic or Latino | 6 (27) | 3 (12) | 7 (8) |
| Not Hispanic or Latino | 16 (73) | 23 (88) | 86 (92) |
| **Age (years), mean (SD)** | 49.9 (14.5) | 64.8 (9.4) | 48.9 (13.8) |
| **Disease type, n (%)** |  |  |  |
| Systemic lupus erythematosus | 16 (73) | 6 (23) | 5 (5) |
| Rheumatoid arthritis | 0 | 20 (77) | 18 (19) |
| Multiple sclerosis | 0 | 0 | 67 (72) |
| Systemic sclerosis | 5 (23) | 0 | 1 (1) |
| Pemphigus | 1 (5) | 0 | 2 (2) |
| **Response to initial COVID-19 vaccination<sup>a</sup>, n (%)** |  |  |  |
| Negative | 9 (41) | 1 (4) | 69 (74) |
| Sub-optimal | 13 (59) | 25 (96) | 24 (26) |
| **Weeks since initial COVID-19 vaccination<sup>b</sup>, mean (SD)** | 24.3 (8.3) | 27.4 (5.7) | 24.9 (6.9) |
| **Prior COVID-19 infection, n (%)** | 1 (5) | 0 | 7 (8) |
| **MMF (mg/day), mean (SD)** | 2325 (832) | NA | NA |
| **MPA (mg/day), mean (SD)** | 2160 (-) | NA | NA |
| **MTX (mg/week), mean (SD)** | NA | 18 (5.4) | NA |
| **Ocrelizumab (mg), mean (SD)** | NA | NA | 585.5 (64.9) |
| **Ofatumumab (mg), mean (SD)** | NA | NA | 20 (-) |
| **Rituximab (mg), mean (SD)** | NA | NA | 960 (148.5) |
| **Baseline CD19 count (cells/uL), mean (SD)** | NA | NA | 11.1 (41.8) |
| **Time from last BCDT (weeks), mean (SD)** | NA | NA | 18.7 (10.3) |

<sup>a</sup>Negative defined as a Roche Elecsys® Anti-SARS-CoV-2 S result $\leq 0.79$ U/mL and sub-optimal defined as a Roche Elecsys® Anti-SARS-CoV-2 S result $> 0.79$ and $\leq 200$ U/mL following the initial COVID-19 vaccine regimen
<sup>b</sup>Defined as weeks since completed initial course of vaccine to informed consent MMF, mycophenolate mofetil; MPA, mycophenolic acid; MTX, methotrexate; BCDT, B cell-depleting therapy

### A third mRNA vaccine dose increases the humoral response against Wuhan-Hu-1 RBD and spike proteins, regardless of withholding medications

At baseline, approximately half of the participants who received an mRNA vaccine were seropositive for Wuhan-Hu-1 RBD in MMF/MPA- (continue, n=5 [50%]; withhold, n=6 [60%]) and MTX-treated (continue, n=7 [54%]; withhold, n=7 [54%]) cohorts (Figure 2, A and B). A third mRNA vaccine dose increased seropositivity to 90% (n=9) and 91% (n=10) in participants who continued MMF/MPA or MTX, respectively, and 90% (n=9) and 100% (n=12) in those who withheld MMF/MPA or MTX; however, the increases in each group were not statistically significant due to the small sample sizes (Figure 2, A and B). Although a third mRNA vaccine dose increased the humoral response in the BCDT-treated cohort from 6% (n=5) to 36% (n=29; p=<0.0001) at 4 weeks, seropositivity was still lower compared to the MMF/MPA- and MTX-treated participants (Figure 2C). Concentrations of antibodies against Wuhan-Hu-1 RBD were significantly higher at 4 weeks post-third mRNA vaccination compared to baseline in all cohorts, and increases in the humoral response did not differ between participants who withheld their medication and those who did not in both the MMF/MPA (p=0.79) and MTX-cohorts (p=0.16) (Figure 2, D-F). Similar humoral response increases were observed against the Wuhan-Hu-1 spike protein and Roche Elecsys® anti-RBD and in participants who received the AD26.COV2.S vaccine (Supplemental Figures 1-3). Humoral responses appeared sustained in the subgroups that remained in the study through 48 weeks, regardless of cohort and treatment withholding (Supplemental Figure 4).

**Figure 2.**
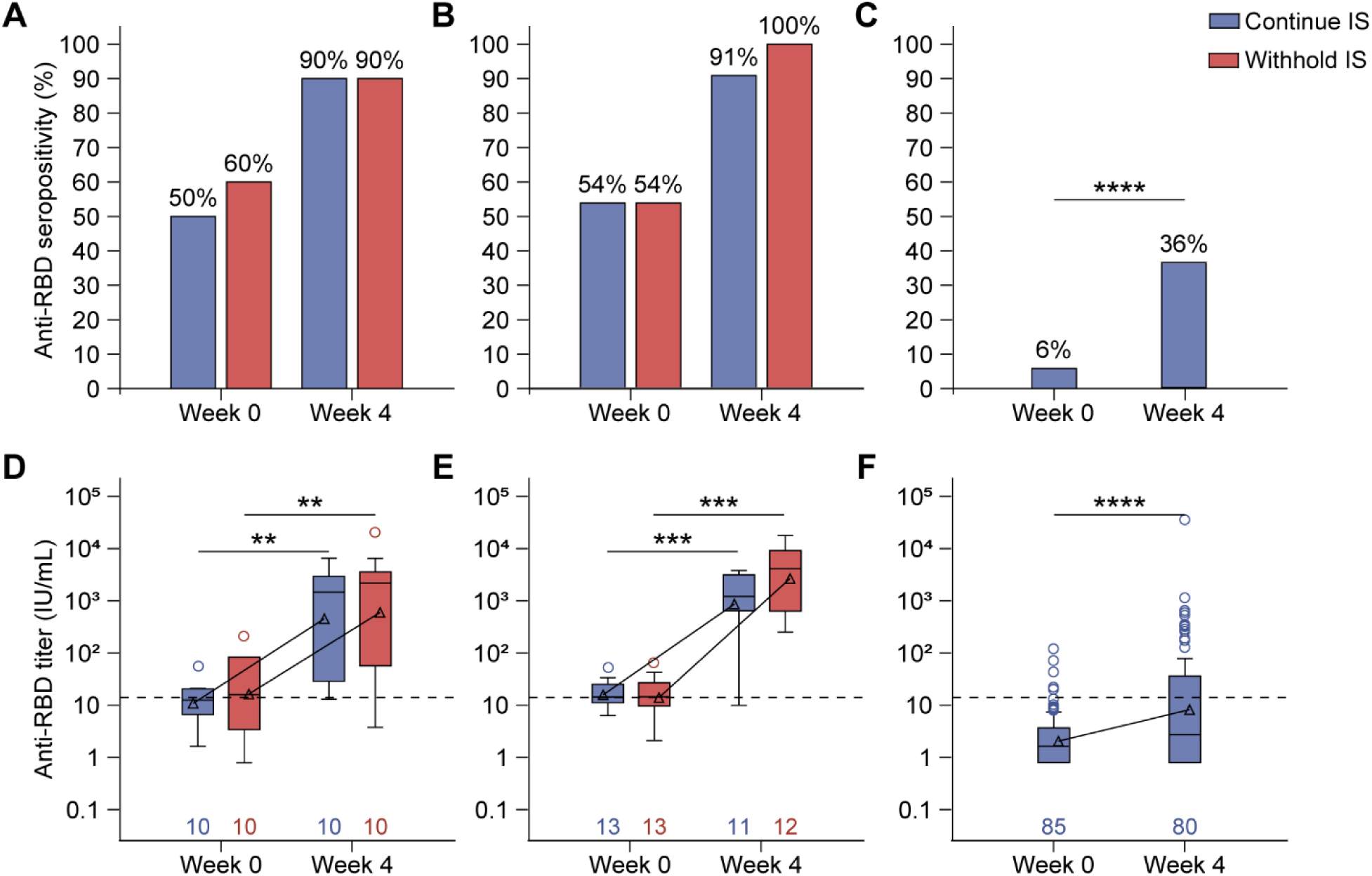
Anti-RBD seropositivity and titers in mycophenolate mofetil/mycophenolic acid (MMF/MPA)-, methotrexate (MTX)-, and B cell-depleting therapy (BCDT)-treated participants who received a third mRNA vaccine. Seropositivity of anti-RBD antibodies at baseline and 4 weeks post-third vaccination in (**A**) MMF/MPA-, (**B**) MTX-, and (**C**) BCDT-treated autoimmune disease patients. Statistical significance was determined using a McNemar test. Concentrations of anti-RBD antibodies at baseline and 4 weeks post-third vaccination in (**D**) MMF/MPA-, (**E**) MTX-, and (**F**) BCDT-treated autoimmune disease patients. Box plots display the interquartile range (IQR), with the line within the box indicating median values. Geometric mean values are represented by triangles and connected across study visits. Whiskers extend to 1.5*IQR, with circles indicating outliers. The dashed line represents the positivity cut-off, and numbers indicate the number of participants analyzed at each time point. The Wilcoxon signed-rank test was used to assess the change in antibody concentration from baseline to Week 4 within each group. There was no significant difference in concentrations between those who continued or withheld immunosuppressants, as determined by a van Elteren test. **p<0.01; ***<0.001; ****p<0.0001.

**Figure 3.**
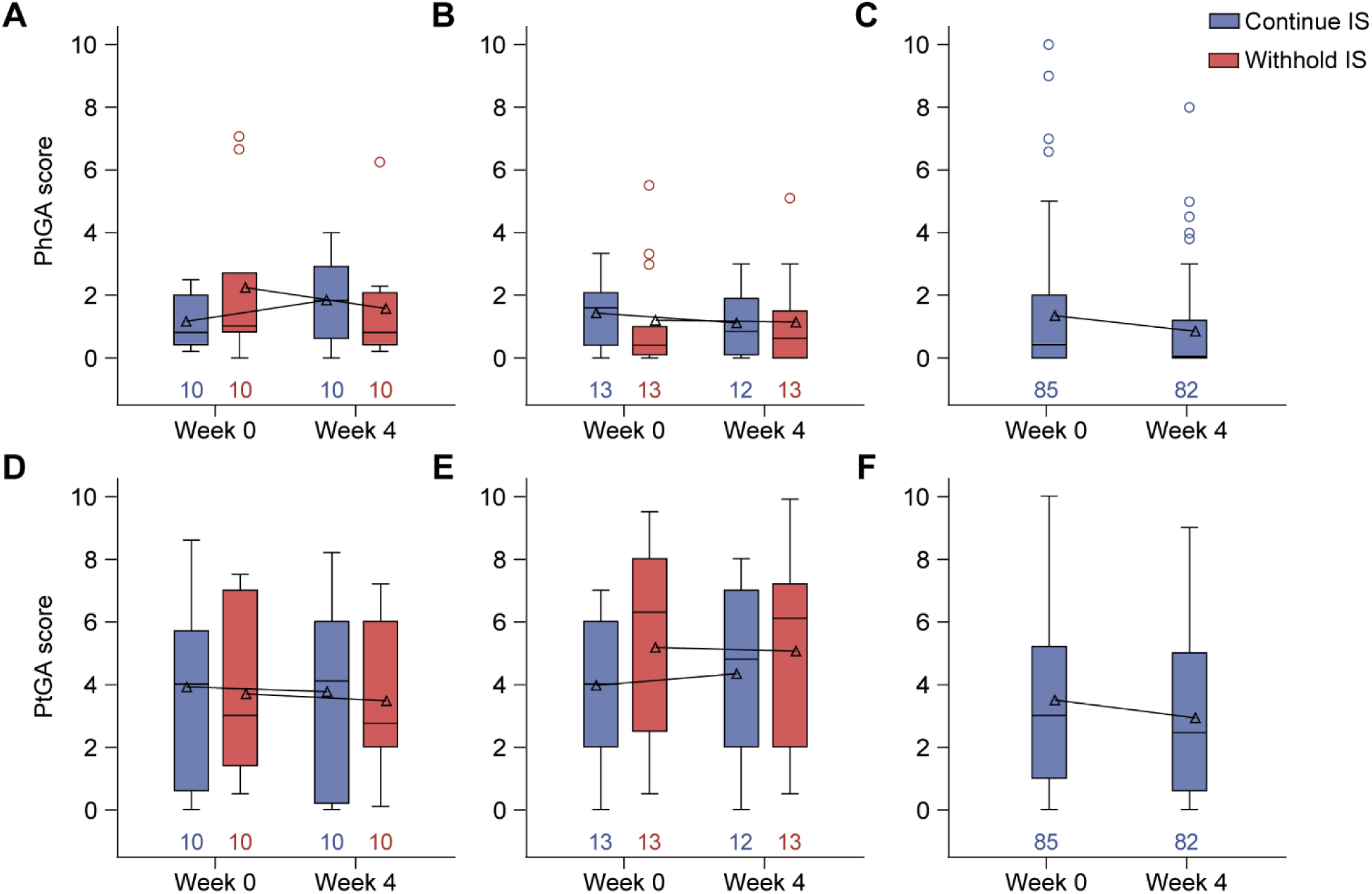
Disease activity in the vaccinated population that received a third mRNA vaccine. Changes in disease activity as measured by the (**A-C**) Physician Global Assessment (PGA) and (**D-F**) Patient Global Assessment (PtGA) at baseline and week 4 post-booster in (**A** and **D**) mycophenolate mofetil/mycophenolic acid (MMF/MPA)-, (**B** and **E**) methotrexate (MTX)-, and (**C** and **F**) B cell-depleting therapy (BCDT)-treated autoimmune disease patients. Box plots display the interquartile range (IQR), with the line within the box indicating median values. Geometric mean values are represented by triangles and connected across study visits. Whiskers extend to 1.5*IQR, with circles indicating outliers. The dashed line represents the positivity cut-off, and numbers indicate the number of participants analyzed at each time point. Change in score from baseline to week 4 did not significantly differ, as determined by the Wilcoxon signed-rank test.

### Antibody response and neutralization across SARS-CoV-2 variants

Next, we examined the effects of a third mRNA vaccine dose on anti-spike antibody concentrations across SARS-CoV-2 variants. Across all cohorts, concentrations of Wuhan-Hu-1 anti-spike antibodies were comparable to those against the Alpha, Beta, and Delta strains (Supplemental Figure 5, A and B). Consistent with the general population (17), participants exhibited lower humoral responses against Omicron spike variants compared to the Wuhan-Hu-1 spike at baseline and 4 weeks post-third mRNA vaccination (Supplemental Figure 5, C and D). Anti-RBD responses to the Wuhan-Hu-1 variant at week 4 post-third mRNA vaccination correlated strongly with Wuhan-Hu-1 live virus neutralization titers (r=0.86; p<0.0001) (Supplemental Figure 6), and fold inhibition of ACE2 binding was similar for the Wuhan-Hu-1, Alpha, Beta, and Delta SARS-CoV-2 strains (Supplemental Figure 7A). However, ACE2 neutralization was significantly reduced against Omicron subvariants BA.1, BA.2, BA.2.12.1, BA.2.75, and BA.5 across all cohorts (Supplemental Figure 7B).

### Disease type and baseline CD19 count are associated with subsequent anti-RBD response

In MMF/MPA- or MTX-treated participants who received an mRNA vaccine, RA patients had the highest anti-RBD concentrations at week 4 post-vaccination compared to those with SLE, SSc, and pemphigus (Supplemental Table 2). In BCDT-treated participants who received an mRNA vaccine, detectable baseline B cell counts, but not the time from the last BCDT dose, was associated with higher anti-RBD concentrations (Supplemental Table 2). Week 4 humoral responses were not associated with the type of mRNA vaccine, medication withholding, gender, age, time since the initial COVID-19 vaccination, prior COVID-19 infection, prednisone use, or type of medication in either cohort.

### COVID-19 infections are mild following a third mRNA vaccine dose

Consistent with a low frequency of infection before additional vaccination (Table 1), seropositivity and titers of antibodies against Wuhan-Hu-1 nucleocapsid were low at baseline and did not increase 4 weeks post-third mRNA vaccination in participants who received an mRNA vaccine (Supplemental Figure 8). COVID-19 infections occurred at a median of 62-110 days, with infections occurring in 30-46% of participants during study participation, with similar frequencies across treatment cohorts (Table 2 and Supplemental Figure 9). All infections (n=7) were deemed mild in the MMF/MPA cohort, but most (6 of 10) were moderate in the MTX cohort (Table 2). Approximately half of the infections were mild in the BCDT cohort. Three hospitalizations occurred (1 in the withhold MTX- and 2 in the BCDT-treated group; Table 2), and none were life-threatening or fatal (grade 4 or 5). Infections were observed across the diseases studied and immunosuppressive therapies (Table 2).

**Table 2.**
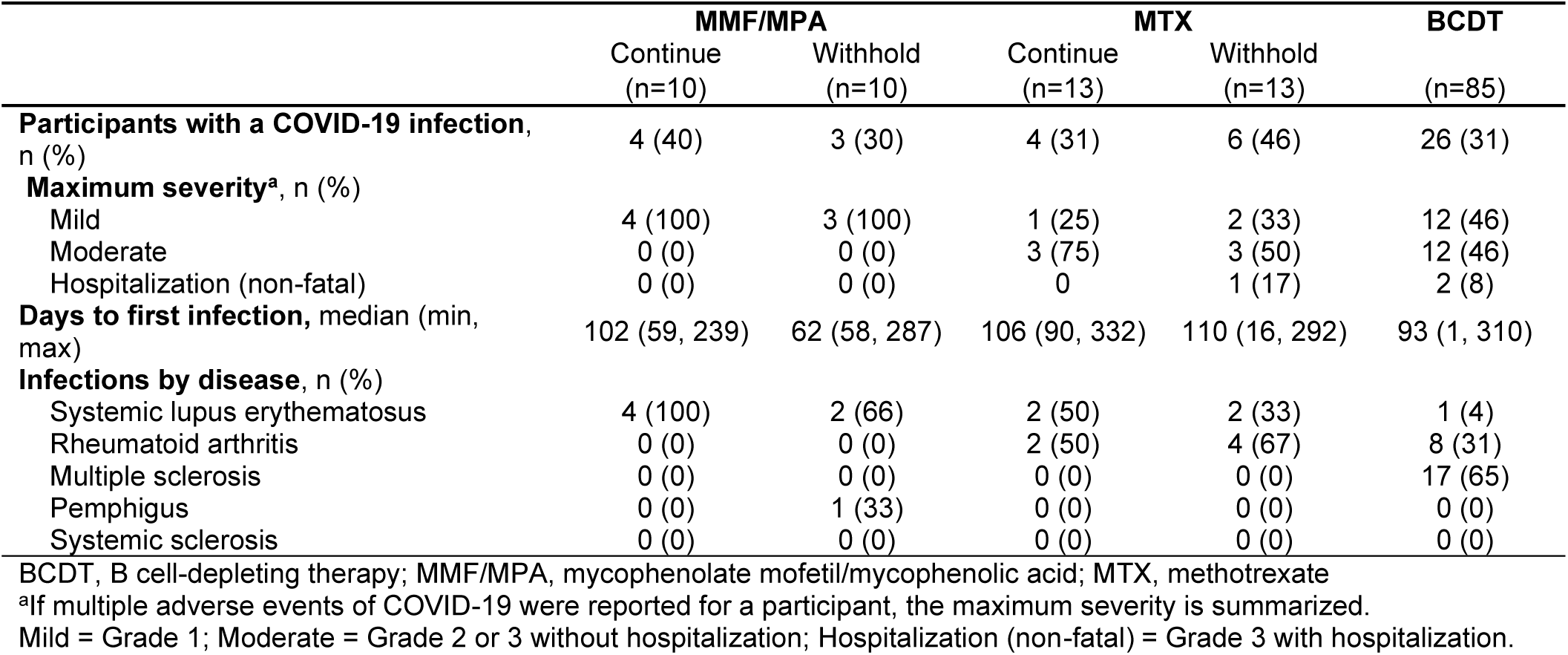
Frequency, severity, and timing of COVID-19 infections throughout the study period (up to 48 weeks post-vaccination) in participants who received an mRNA vaccine.

### A third mRNA vaccine dose is safe in autoimmune disease patients and does not affect disease activity

Within 7 days of additional vaccination, local and systemic reactions were predominantly mild to moderate across all cohorts and vaccines (Supplemental Figure 10). Throughout the study, 39-46% of participants who received an mRNA vaccine reported AEs, 6 of which (all BCDT-treated) were related to the vaccine (Supplemental Table 3). Furthermore, most AEs were grade 1 or 2, with 1 participant (8%) in the withhold MTX cohort and 9 participants (11%) in the BCDT-treated cohort experiencing a serious AE (SAE), of which all were grade 3, and only 1 was categorized as vaccine-related (Supplemental Table 3). No grade 4 or grade 5 AEs occurred. In the BCDT-treated cohort, 1 (1%) participant experienced a medically attended AE (MAAE), and 7 participants (8%) experienced a new-onset chronic medical condition (NOCMC), none of which was a new autoimmune diagnosis (Supplemental Table 3). No participants experienced myocarditis or pericarditis during the study period (Supplemental Table 3). Participants who received the AD26.COV2.S vaccine did not experience vaccine-related AEs (Supplemental Table 4).

In all cohorts, the Physician’s Global Assessment and Patient’s Global Assessment of disease activity were similar at baseline and 4 weeks post-third mRNA vaccine dose, regardless of whether medications were withheld (Figure 3). Similarly, most participants had no change based on the Clinical Global Impression of Change and Patient Global Impression of Change scales 4 weeks post-third mRNA vaccination, with some participants who withheld MMF/MPA or MTX reporting that their symptoms improved. (Supplemental Figure 11). While post-vaccine disease flares occurred in 17 (13%) participants, severe flares were rare, occurring in only 1 RA (BCDT) and 1 pemphigus (MMF/MPA withheld) patient (Supplemental Table 5).

## Discussion

In this randomized, multi-site, open-label clinical trial, we found that mRNA booster vaccination enhances humoral responses to the SARS-CoV-2 RBD and spike proteins, regardless of whether immunosuppressive medication was continued or withheld. Although lower than responses against the WT spike, humoral responses were also increased for other SARS-CoV-2 variants, including Omicron. Breakthrough infections occurred, but they were typically mild in all treatment groups. Booster vaccination was safe in autoimmune disease patients, with no vaccine-related SAEs or changes in disease activity. Flare rates were similar across treatment groups, and severe flares were rare. Despite a limited sample size, we found similar trends in the humoral response in participants treated with the AD26.COV2.S vaccine.

We found that mRNA booster vaccination increases humoral responses to SARS-CoV-2 in autoimmune disease patients taking MTX, with similar concentrations and seropositivity in those who withheld MTX and those who did not. This finding contrasts with the VROOM clinical trial, which found a 2-fold increase in anti-RBD titers in patients who suspended MTX treatment for 2 weeks following mRNA or Oxford-AstraZeneca ChAdOx1 nCoV-19 (AZD1222) booster vaccination compared to those who did not (15). As booster vaccination with AZD1222 appeared to have a greater effect on the antibody response compared to BNT162b2 and mRNA-1273 in the VROOM clinical trial (15), we may not have seen an improvement in the antibody response as we only analyzed mRNA vaccines. Supporting this hypothesis, the RESCUE 2 trial demonstrated a trend towards lower antibody levels in immune-mediated inflammatory disease patients who continued conventional synthetic disease-modifying antirheumatic drug regimens, the majority of which contained MTX, compared to those who withheld their medication or compared to healthy controls following booster vaccination with AZD1222; however, withholding these drugs did not impact the humoral response in patients receiving mRNA booster vaccination (18). Despite increased antibody titers, the VROOM clinical trial observed mild SARS-CoV-2 infections with similar frequencies of throughout the study, consistent with our findings (15).

Results from observational studies are conflicting on the effects of MMF/MPA withholding around the time of initial and booster vaccination, and clinical trials are lacking in autoimmune disease patients (16,19). However, a clinical trial in kidney transplant recipients who withheld MMF/MPA for 1 week before and 1 week after the 3^rd^ or 4^th^ dose of the mRNA vaccine found no difference in antibody concentrations or neutralizing activity compared to those who continued MMF/MPA (20). To our knowledge, our study is the first randomized clinical trial of MMF/MPA withdrawal in autoimmune disease patients.

Although anti-RBD seropositivity increased from 6% to 36% 4 weeks post-third mRNA vaccination, it remained lower compared to MMF/MPA- and MTX-treated participants. This finding aligns with a European clinical trial in which only 32% of 28 rituximab-treated patients seroconverted following a third mRNA vaccine (21). As previously described (22,23), baseline CD19 counts were associated with subsequent seropositivity against RBD, suggesting that waiting until B cells have repopulated before vaccination could improve vaccine responses in these patients. Despite the lower antibody responses, breakthrough infection rates and severity were similar across the treatment cohorts. Robust SARS-CoV-2 specific CD4 and CD8 T cell responses are observed in BCDT-treated patients, similar to healthy controls, especially among those who did not develop anti-RBD antibodies (22). As SARS-CoV-2-specific T cells are associated with better COVID-19 outcomes (24,25), T cell responses may provide an alternative protective mechanism in these patients, compensating for lower humoral responses.

Consistent with observations in the general population (17), humoral responses to and neutralization of the Omicron variant were significantly lower compared to earlier SARS-CoV-2 strains. However, although our study occurred during the Omicron outbreak, subsequent infections were mild, with only 3 (2% of participants) non-fatal hospitalizations, reflecting a hospitalization risk comparable to that seen in the general population (26). These findings suggest that additional vaccination prevents severe outcomes against newer SARS-CoV-2 variants, even with reduced humoral responses. Since T cells are less impacted by spike protein mutations and continue to recognize mutant SARS-CoV-2 variants (27,28), they may play a compensatory role in protecting against severe disease. Nevertheless, additional or variant-adapted vaccine doses would likely enhance immune protection, especially in BCDT-treated patients, who generally exhibit lower humoral responses.

We observed minimal adverse events and no significant impact on disease activity or flare following additional vaccination, reinforcing the safety of booster doses in autoimmune disease patients on immunosuppressive therapies (29–31). Notably, disease flares were minimal 4 weeks post-third mRNA vaccination in patients who temporarily withheld MMF/MPA or MTX around the time of vaccination, with only 1 severe flare occurring among the 11 participants who withheld these medications. In the VROOM clinical trial, self-reported flares were initially higher in participants who withheld MTX compared to those who continued MTX 4 and 12 weeks post-booster; however, these differences resolved within 26 weeks, with most flares not requiring medical attention (14,15). Further supporting autoimmune disease stability, autoantibody levels and type I IFN signature gene expression remained stable after both the second and third mRNA vaccine doses (31,32). Together, these findings support the safety of additional mRNA vaccination in autoimmune disease patients on immunosuppressive therapies, even among those who withheld medications.

In conclusion, additional COVID-19 vaccination is both effective and safe for autoimmune disease patients treated with immunosuppressive therapies, with no clear observed benefit in humoral response or safety outcomes from withholding these medications around the time of vaccination. Thus, findings from our study support the continued use of COVID-19 vaccines in this population without the need for adjusting immunosuppressive treatments. However, further studies are warranted to explore strategies to improve humoral responses in patients who maintain suboptimal responses, including the potential benefits of additional vaccine doses, heterologous booster vaccines, and variant-specific vaccines.

## Methods

### Trial design and sex as a biological variable

The COVID-19 Booster Vaccine in Autoimmune Disease Non-Responders (NCT05000216) study was a randomized, multi-site, adaptive, open-label clinical trial that included male and female participants. This study consisted of two stages: Stage 1 participants received an additional homologous vaccine dose, and Stage 2 participants received an additional heterologous vaccine dose; only Stage 1 results are reported here. Outcomes in male and female participants are reported together. The study was funded by the Autoimmunity Centers of Excellence Network and sponsored by the National Institute of Allergy and Infectious Diseases at the NIH (NIAID/NIH). Study vaccines were provided by Health and Human Services Coordination Operations and Response Element, formerly Countermeasures Acceleration Group and Operation Warp Speed, and distributed to each clinical site by the Division of Allergy, Immunology, and Transplantation/NIAID’s Clinical Products Center.

### Eligibility criteria

Eligible participants were aged 18 years and older and met at least one set of 2019 American College of Rheumatology (ACR)/European Alliance of Associations for Rheumatology (EULAR) (33) or 2012 SLICC classification criteria (34) for SLE, 2010 ACR/EULAR classification criteria for RA (35), 2013 ACR/EULAR classification criteria for SSc (36), 2017 McDonald criteria for MS (37), or the international consensus criteria for pemphigus (38). Participants were also required to currently be taking MMF (minimum of 1000 mg/day for at least 8 weeks) or MPA (minimum of 720 mg/day for at least 8 weeks), MTX (minimum of 7.5 mg/week for at least 8 weeks), or an anti-CD20 or anti-CD19 BCDT in the previous 18 months. Participants had to have received two primary doses of the BNT162b2 (Pfizer–BioNTech) or mRNA-1273 (Moderna) vaccine or one dose of the AD26.COV2.S (Janssen) vaccine at least 4 weeks and no more than 52 weeks prior to screening and have had a negative (<0.79 U/mL, Roche Elecsys® Anti-SARS-CoV-2 S) or suboptimal (≤200 U/mL, Roche Elecsys® Anti-SARS-CoV-2 S) response to the initial COVID-19 vaccine regimen. Exclusion criteria included history of severe allergic reaction to the initial COVID-19 vaccine regimen or any component of the COVID-19 vaccine, female participants planning a pregnancy during the trial, receipt of a COVID-19 booster vaccine, receipt of anti-SARS-CoV-2 monoclonal antibodies or plasma products within 90 days of screening, history of thrombosis in the previous 12 months, history of COVID infection within 30 days of screening, and ongoing dialysis or history of a solid organ transplant. Subjects with active disease resulting in an inability to withhold IS therapy or necessitating increased doses or the addition of new IS in the MTX or MMF/MPA arms were also excluded.

### Sampling and randomization of subjects

Participants were assigned to cohorts defined by their IS medication (MMF/MPA-, MTX-, or BCDT-treated) and their original COVID-19 vaccine regimen. If a participant had received BCDT within the previous 18 months and was also on MMF/MPA or MTX, they were included in the BCDT cohort. Participants in the MMF/MPA- and MTX-treated cohorts were randomized one-to-one within the cohorts to continue or withhold their IS medication. Study site staff randomized participants through a centralized, web-based, password-protected randomization system. Randomization was performed using a permuted block design stratified by disease type.

### Trial procedure

All participants received an additional homologous dose of COVID-19 vaccine at the baseline visit. In the withhold groups, MMF/MPA was withheld for 3 days before and 10 days after the booster vaccine, while MTX was withheld for at least 7 days before and at least 7 days after the booster vaccine, but for no more than 21 days total. Participants randomized to continue MMF/MPA or MTX were required to maintain a stable dose during the study period. Participants in the BCDT cohort continued to take BCDT without pre-specified alterations in schedule and dosing. All participants were asked to continue taking stable doses of any additional IS.

### Antibody responses

Antibody responses to Wuhan-Hu-1 full-length spike, RBD, and N proteins were measured using the V-PLEX SARS-CoV-2 384 Panel 1 IgG kit (Cat# K25392U; Meso Scale Diagnostics, LCC. Rockville, USA) as previously described (39). Seropositivity was defined as concentrations above the positive threshold value determined during assay validation with pre-2019 samples (spike 10.842 IU/mL; RBD 14.086 IU/mL) (39). Antibody responses to spike proteins of different SARS-CoV-2 variants were measured as AU/mL relative to a reference standard using the V-PLEX SARS-CoV-2 Key Variant Spike Panel 1 IgG kit (Cat# K15651U; Meso Scale Diagnostics, LCC. Rockville, USA) as previously described (40).

### Neutralization

Plasma (1:40 initial dilution) was serially diluted 1:2 across a 96-well plate and mixed with enough virus (isolate USA-WA1/2020) to yield a final multiplicity of infection of 0.01. After incubating the antibody virus mixture for 1 h at 37°C, the virus mixture was transferred to a 96-well plate containing VERO E6 cells seeded at 10,000 cells per well. SARS-CoV-2 activity was determined 96 hours after infection by visually observing cytopathic effects. The antibody dilution at which virus-positive wells were observed was recorded.

The V-PLEX ACE2 Neutralization Kit (Cat# K15654U; Meso Scale Diagnostics, LCC. Rockville, USA) was used to detect surrogate neutralizing antibodies across SARS-CoV-2 variants as previously described (40).

### Outcomes

The primary outcome was the increase in anti-RBD at 4 weeks post-additional homologous COVID-19 vaccination. Secondary outcomes related to antibody response included seropositivity and/or antibody titers for other SARS-CoV-2 antigens and variants, neutralization, and longitudinal changes in antibody responses. Additional secondary outcomes included changes in global disease activity, as measured by the Clinical Global Impression of Change (CGI-C) (41) and the Physician Global Assessment (PGA) (42), and changes in patient-reported outcomes, as measured by the Participant-Reported Outcomes Measurement Information System (PROMIS) 29 (43), Patient Global Assessment (PtGA) (42), and Patient Global Impression of Change (PGI-C) (44) across all 5 diseases. Changes in disease-specific activity were also measured using the following disease-specific assessments: Hybrid-SLEDAI and Thanou modified SELENA-SLEDAI Flare Index (45) for SLE participants, the Disease Activity Score-28 CRP (46) for RA, the modified Rodnan Skin Score (mRSS) (47) for SSc, the Kurtzke Expanded Disability Status Scale (EDSS) (48) for MS, and the Pemphigus Disease Pemphigus Disease Area Index (PDAI) (49) for pemphigus.

Safety outcomes included the proportion of participants who experienced any Grade 1 or higher AE related to the COVID-19 vaccine, the incidence of solicited AEs that were local and systemic events of reactogenicity, and any SA, MAAE, NOCMC, or COVID-19 infection. Adverse events of special interest included the incidence of myocarditis and pericarditis. Solicited events of reactogenicity were reported from day 1 through day 7, AEs were reported from day 1 through day 28, and SAEs, MAAEs, NOCMCs, and COVID-19 infections were reported from day 1 through the end of study participation. AEs were graded according to the Common Terminology Criteria for Adverse Events, version 5.0. COVID-19 infections were confirmed by molecular COVID-19 testing. Visits to assess endpoints occurred at baseline (week 0) and weeks 4, 12, 24, 36, and 48.

### Statistics

In each arm, up to 60 participants were to be randomized or allocated. The number of participants with Roche Elecsys® Anti-SARS-CoV-2 S results ≤50 U/mL and with results >50 U/mL and ≤200 U/mL was capped at 40 participants per group. Sample sizes were defined to test whether responses to the additional vaccine dose were greater than an ineffective rate of seroprotection that was less than or equal to 0.25 (null hypothesis). Under this assumption, a sample size of 33 evaluable participants per arm achieved at least 90% power to reject the null hypothesis when the true response rate is 0.5 or greater, using a one-sided exact test with alpha = 0.05. A sample size of 40 participants per arm was originally planned to account for a 20% dropout rate. Later versions of the protocol amended the sample size to 60 participants per arm to account for expansions in the Roche Elecsys eligibility criteria.

A threshold for achieving a protective antibody response was not established for the Meso Scale Diagnostics SARS-CoV-2 assays, so vaccine response is evaluated as a seropositivity rate and a continuous measure of antibody concentrations. Descriptive statistics included median, minimum, maximum, interquartile range, geometric mean, coefficient of variation of the geometric mean, and 95% confidence intervals for the Wuhan-Hu-1 spike and RBD proteins. For concentrations below the lower limit of detection (LLOD), numeric values equivalent to LLOD/2 were assigned; concentrations above the upper limit of quantification were reported without adjustments. Antibody results collected after a documented COVID-19 infection, monoclonal antibody use, or a COVID-19 vaccine given off-study were excluded from the analyses. Seropositivity, defined as having a result greater than the positive threshold value, was also summarized for the Wuhan-Hu-1 spike and RBD proteins. Descriptive statistics were reported in the vaccinated population (all participants who received an additional vaccination) within each group (defined by cohort, vaccine, and IS treatment plan) and in the combined groups that received an mRNA vaccine.

Categorical variables were summarized with frequencies and proportions. The Wilcoxon signed rank test assessed changes in antibody concentrations within arms and subgroups over time. Comparisons across arms and predictors of response were assessed using the Wilcoxon rank sum test and stratified van Elteren tests. Changes in the seropositivity rate over time were evaluated using the McNemar test. The correlation between immune response endpoints is explored in the vaccinated population using Spearman’s rank correlation coefficients. The cumulative incidence of breakthrough COVID-19 infections was calculated using the Kaplan-Meier product limit estimator using Greenwood’s formula for standard error. Statistical analyses were performed using SAS 9.3 or higher (SAS Institute; Cary, NC). P-values were not adjusted for multiple comparisons. P-values less than 0.05 were considered statistically significant.

### Study approval

Participants were recruited from 19 US centers (Supplemental Table 6), and all provided written informed consent before participation. The study was approved by a central institutional review board (Advarra, Columbia, MD), reviewed by institutional review boards at each site, and conducted following the Declaration of Helsinki. An independent Data and Safety Monitoring Board reviewed the progress of the study and safety data twice per year.

### Data availability

All data are accessible from ImmPort (www.immport.org).

## Author contributions

MM, AP, WTB, EG, JM, ABO, DK, JAJ designed the research and KS, JMG, SM, JW, GD, MC, RC, SN, BCL, LS, AM conducted experiments. MM, AP, JAC, ZSW, AK, JAS, SL, AS, JW, GD, MC, RC, AHJK, DLK, FK, ADA, KS, JMG, GP, YMD, SM, SM, MAS, SDH, MV, SW, KY, ST, RC, SN, BCL, LS, AM, WTB, ASP, AB, DK, and JAJ acquired data and WTB, AP, CAW, MM, ABO, DK, JAJ analyzed data. All authors wrote and/or reviewed the manuscript.

## Acknowledgments

We thank the ACV01 protocol development team, site clinical research teams, and participants. See Supplemental Acknowledgements for details on the ACV01 Study Team. The study was supported by awards from the Autoimmunity Centers of Excellence, a research network funded by the National Institute of Allergy and Infectious Diseases (NIAID/NIH) (U19AI144306, U19AI082714, U19AI110483, UM1AI110494, UM1AI144292, UM1AI144295, UM1AI144288, UM1AI144298, UM1AI110557) and to Rho, the statistical and clinical coordinating center (UM2AI117870; U19AI110483-08S4; 75N93022C00003).

