## Supplemental for "Prospective SARS-CoV-2 Booster Vaccination in Immunosuppressant-Treated Systemic Autoimmune Disease Patients in a Randomized Controlled Trial"

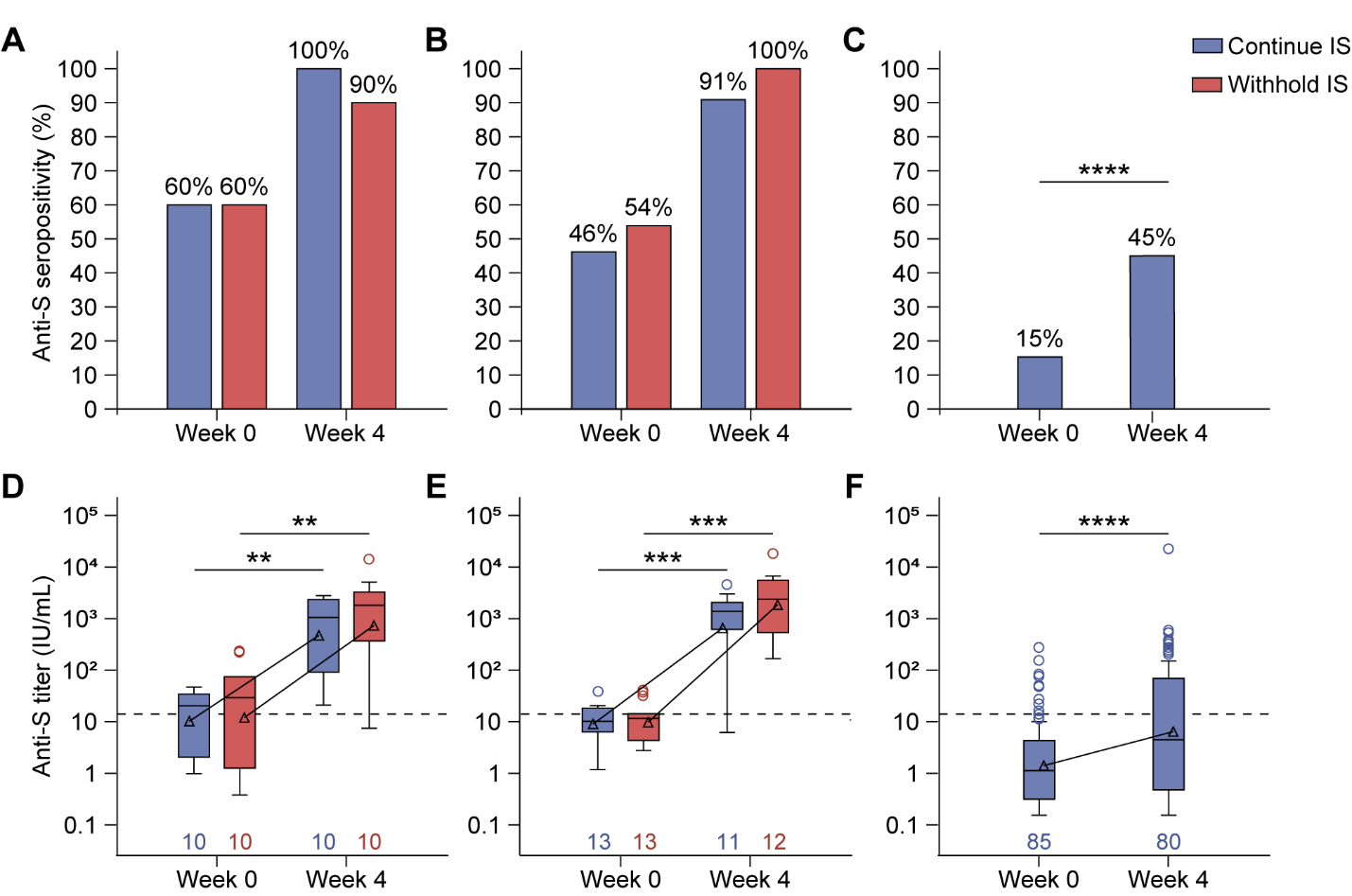
**Supplemental Figure 1. Anti-Spike (S) seropositivity and titers in mycophenolate mofetil/mycophenolic acid (MMF/MPA)-, methotrexate (MTX)-, and B cell-depleting therapy (BCDT)-treated participants who received a third mRNA vaccine.** Seropositivity of anti-S antibodies at baseline and 4 weeks post-third vaccination in (**A**) MMF/MPA-, (**B**) MTX-, and (**C**) BCDT-treated autoimmune disease patients. Statistical significance was determined using a McNemar test. Concentrations of anti-RBD antibodies at baseline and 4 weeks Box plots display the interquartile range (IQR), with the line within the box indicating median values. Geometric mean values are represented by triangles and connected across study visits. Whiskers extend to 1.5*IQR, with circles indicating outliers. The dashed line represents the positivity cut-off, and numbers indicate the number of participants analyzed at each time point. The Wilcoxon signed-rank test was used to assess the change in antibody concentration from baseline to Week 4 within each group. No significant difference in concentrations in those who continued or withheld immunosuppressants using a van Elteren test. **p<0.01; ***<0.001; ****p<0.0001.

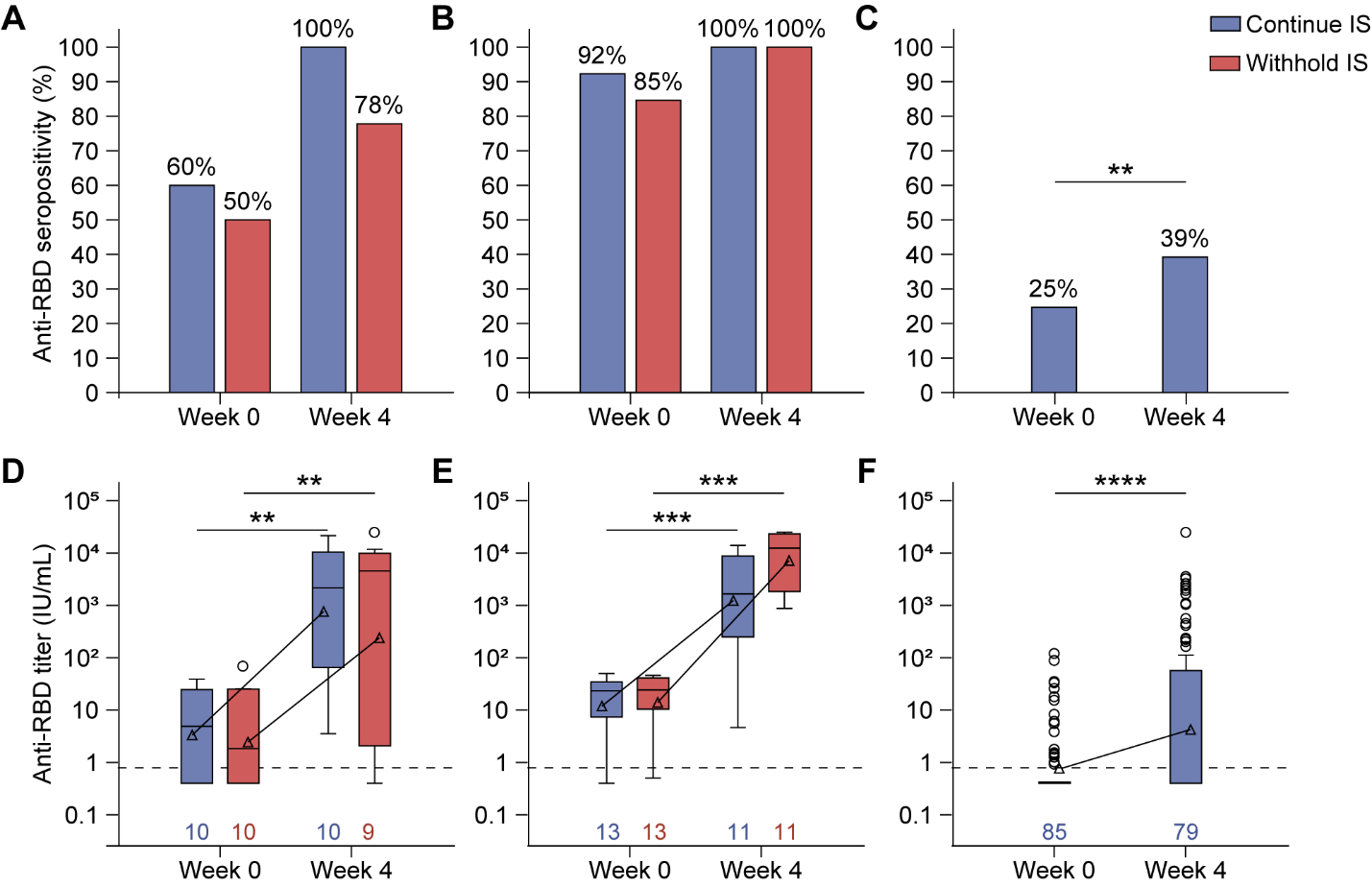

**Supplemental Figure 2.** **Roche Elecsys® anti-spike seropositivity and titers in mycophenolate mofetil/mycophenolic acid (MMF/MPA)-, methotrexate (MTX)-, and B cell-depleting therapy (BCDT)-treated participants who received a third mRNA vaccine.** Seropositivity of Roche Elecsys® anti-RBD antibodies at baseline and 4 weeks post-third vaccination in (**A**) MMF/MPA-, (**B**) MTX-, and (**C**) BCDT-treated autoimmune disease patients. Statistical significance was determined using a McNemar test. Concentrations of anti-RBD antibodies at baseline and 4 weeks post-third vaccination in (**D**) MMF/MPA-, (**E**) MTX-, and (**F**) BCDT-treated autoimmune disease patients. Box plots display the interquartile range (IQR), with the line within the box indicating median values. Geometric mean values are represented by triangles and connected across study visits. Whiskers extend to 1.5*IQR, with circles indicating outliers. The dashed line represents the positivity cut-off, and numbers indicate the number of participants analyzed at each time point. The Wilcoxon signed-rank test was used to assess the change in antibody concentration from baseline to Week 4 within each group. No significant difference in concentrations in those who continued or withheld immunosuppressants using a van Elteren test. **p<0.01; ***<0.001; ****p<0.0001.

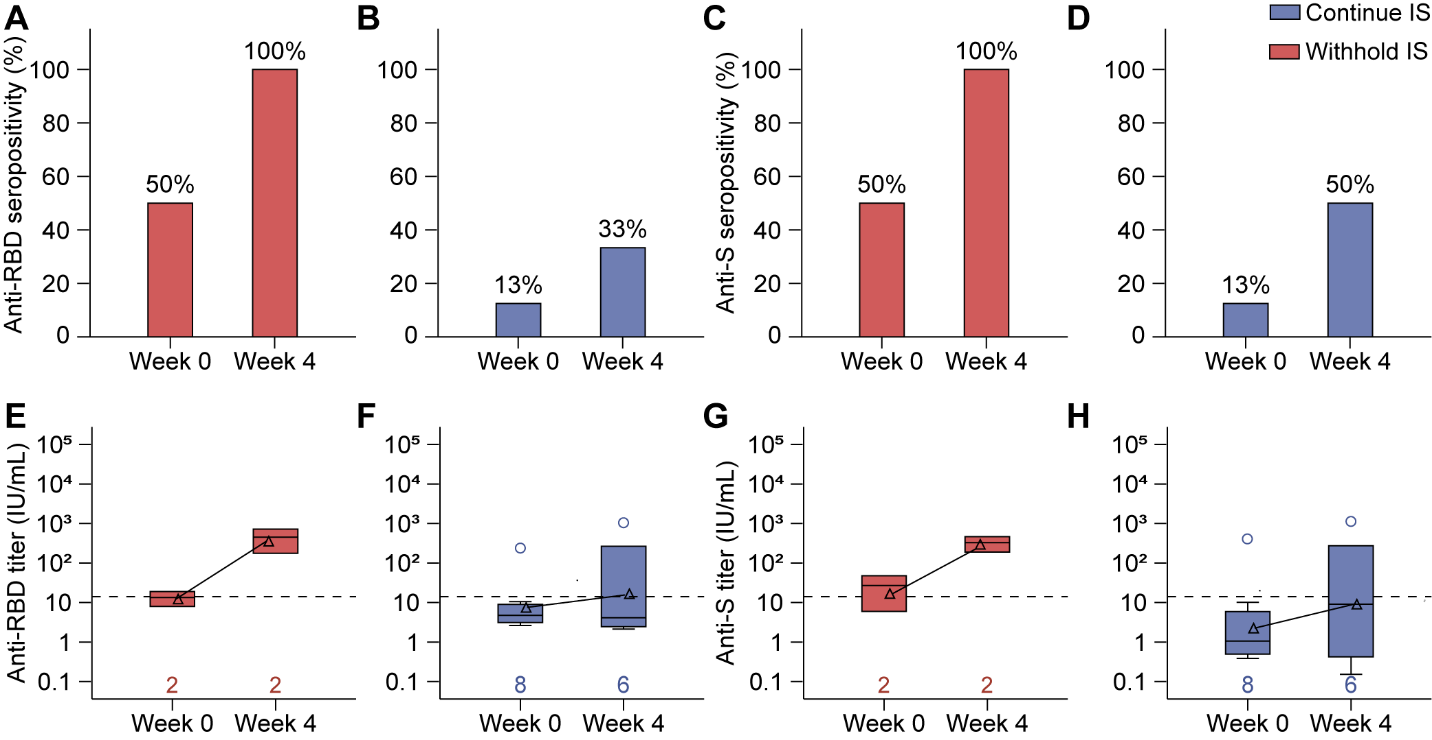

**Supplemental Figure 3. Humoral response in participants who received a second AD26.COV2.S** **vaccine.** (**A-D**) Seropositivity and (**E-F**) concentration of (**A-B** and **E-F**) anti-RBD and (**C-D** and **G-H**) anti-Spike (S) antibodies at weeks 0 and 4 post-second AD26.COV2.S vaccination in (**A, C, E,** and **G**) mycophenolate mofetil/mycophenolic acid (MMF/MPA)-treated autoimmune disease patients who withdrew immunosuppressant use and (**B, D, F,** and **H**) B cell-depleting therapy (BCDT)-treated patients. Box plots display the interquartile range (IQR), with the line within the box indicating median values. Geometric mean values are represented by triangles and connected across study visits. Whiskers extend to 1.5*IQR, with circles indicating outliers. The dashed line represents the positivity cut-off, and numbers indicate the number of participants analyzed at each time point.

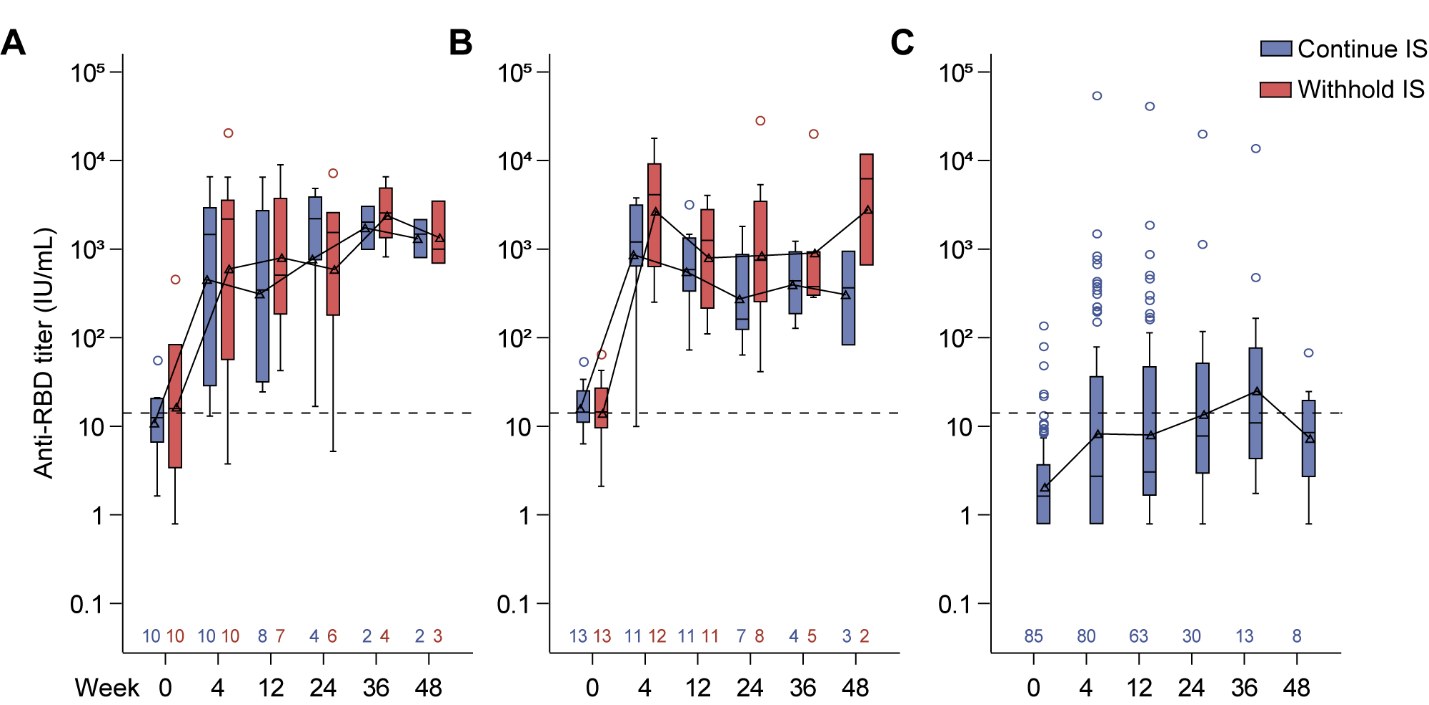
**Supplemental Figure 4. Longitudinal anti-RBD responses post-third mRNA vaccination in mycophenolate mofetil/mycophenolic acid (MMF/MPA)-, methotrexate (MTX)-, and B cell-depleting therapy (BCDT)-treated participants.** Anti-RBD antibodies were measured at baseline and 4, 12, 24, 36, and 48 weeks post-third vaccination in (**A**) MMF/MPA-, (**B**) MTX-, and (**C**) BCDT-treated autoimmune disease patients. Box plots display the interquartile range (IQR), with the line within the box indicating median values. Geometric mean values are represented by triangles and connected across study visits. Whiskers extend to 1.5*IQR, with circles indicating outliers. The dashed line represents the positivity cut-off, and numbers indicate the number of participants analyzed at each time point; samples collected after a documented COVID-19 infection, monoclonal antibody use, or a COVID-19 vaccination given off-study are excluded from the analyses. Some participants with low or sub-optimal responses rolled over to Stage 2 of the study prior to completing Stage 1.

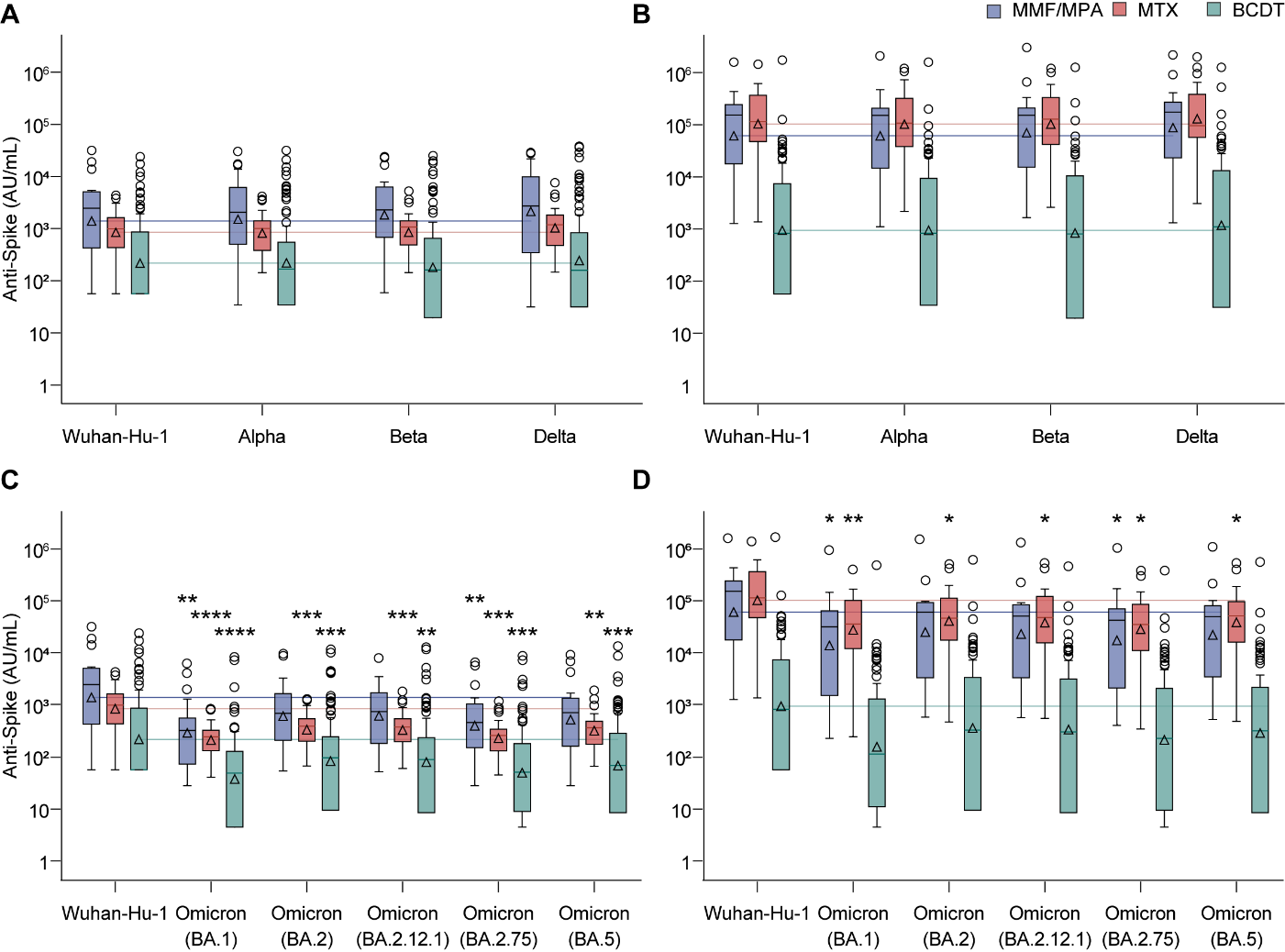

**Supplemental Figure 5. Anti-Spike (S) humoral responses across SARS-CoV-2 variants.** Antibody titers against the (**A-B**) Alpha, Beta, and Delta and (**C-D**) Omicron BA1, BA2, BA2.12.1, BA2.75, and BA5 S proteins relative to Wuhan-Hu-1 at (**A** and **C**) baseline and (**B** and **D**) week 4 post-third mRNA booster vaccination. Box plots display the interquartile range (IQR), with the line within the box indicating median values. Geometric mean values are represented by triangles. Whiskers extend to 1.5*IQR, with circles indicating outliers. Horizontal lines indicate the geometric mean for the Wuhan-Hu-1 Spike variant by cohort for each visit. Statistical significance was determined using the stratified van Elteren test. *p<0.05, **p<0.01 relative to the Wuhan-Hu-1 S protein for each cohort.

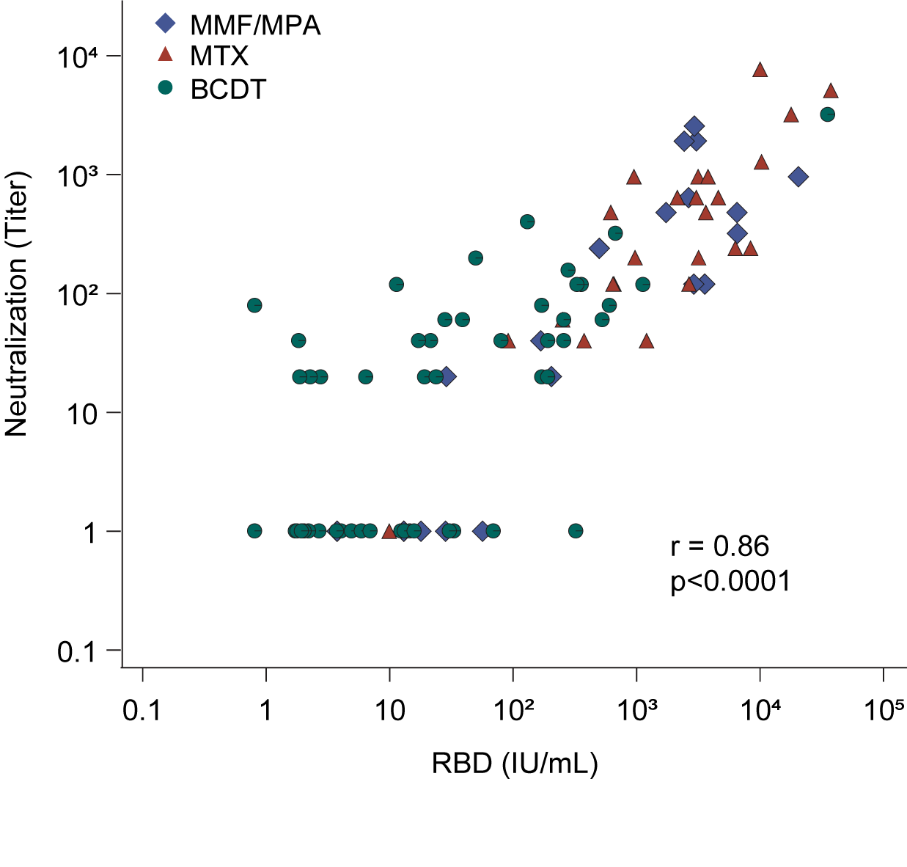

**Supplemental Figure 6.** **Correlation** **of week 4 anti-RBD and neutralization titers in participants who received a third mRNA vaccine.** Correlation between week 4 anti-RBD antibody titers and neutralization titers against the USA-WA1/2020 isolate. Correlation was assessed using Spearman’s rank correlation coefficients.

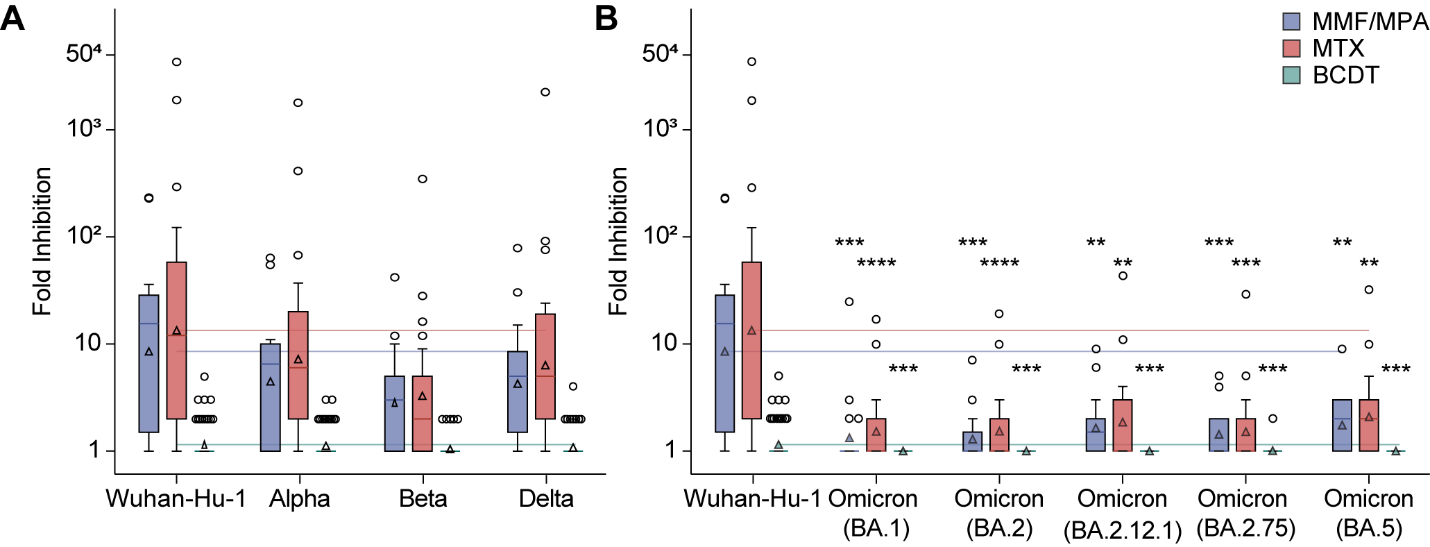

**Supplemental Figure 7. ACE2 neutralization across SARS-CoV-2 variants.** Fold inhibition of ACE2 binding against the (**A**) Alpha, Beta, and Delta and (**B**) Omicron BA.1, BA.2, BA.2.12.1, BA.2.75, and BA.5 SARS-CoV-2 variants relative to Wuhan-Hu-1 at week 4 post-third mRNA booster vaccination. Box plots display the interquartile range (IQR), with the line within the box indicating median values. Geometric mean values are represented by triangles and connected across study visits. Whiskers extend to 1.5*IQR, with circles indicating outliers. Horizontal reference lines indicate the geometric mean for the Wuhan-Hu-1 variant by cohort for each visit. Statistical significance was determined using the stratified van Elteren test. **p<0.01, ***p<0.001, ****p<0.0001 relative to Wuhan-Hu-1 for each cohort.

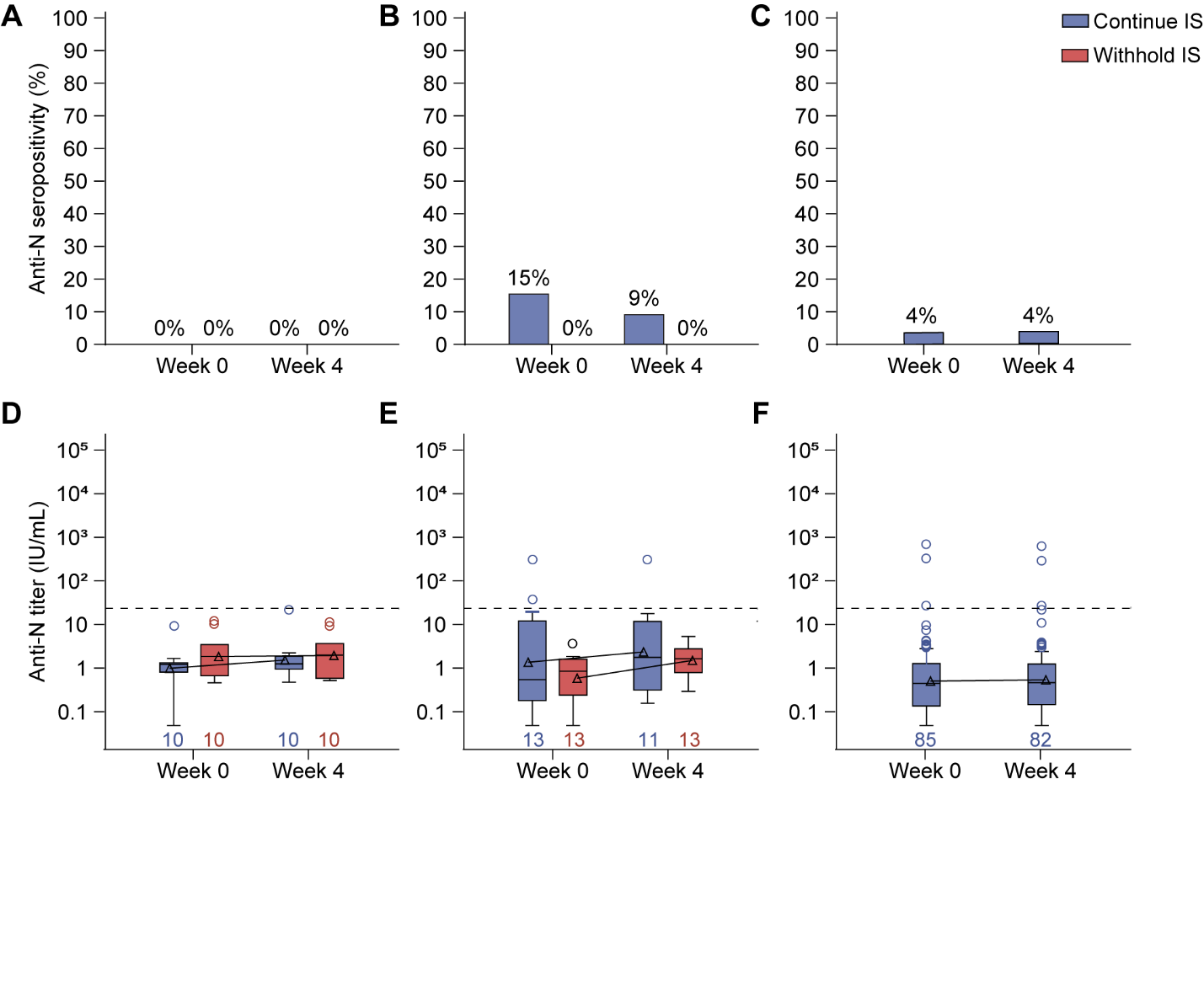
**Supplemental Figure 8. Anti-N-protein antibodies in participants who received a third mRNA vaccine.** Seropositivity of anti-N-protein antibodies at baseline and 4 weeks post-third mRNA vaccination in (**A**) mycophenolate mofetil/mycophenolic acid (MMF/MPA)-, (**B**) methotrexate (MTX)-, and (**C**) B cell-depleting therapy (BCDT)-treated autoimmune disease patients. Concentrations of anti-RBD antibodies at baseline and 4 weeks post-third mRNA vaccination in (**D**) MMF/MPA-, (**E**) MTX-, and (**F**) BCDT-treated autoimmune disease patients. Box plots display the interquartile range (IQR), with the line within the box indicating median values. Geometric mean values are represented by triangles and connected across study visits. Whiskers extend to 1.5*IQR, with circles indicating outliers. The dashed line represents the positivity cut-off, and numbers indicate the number of participants analyzed at each time point.

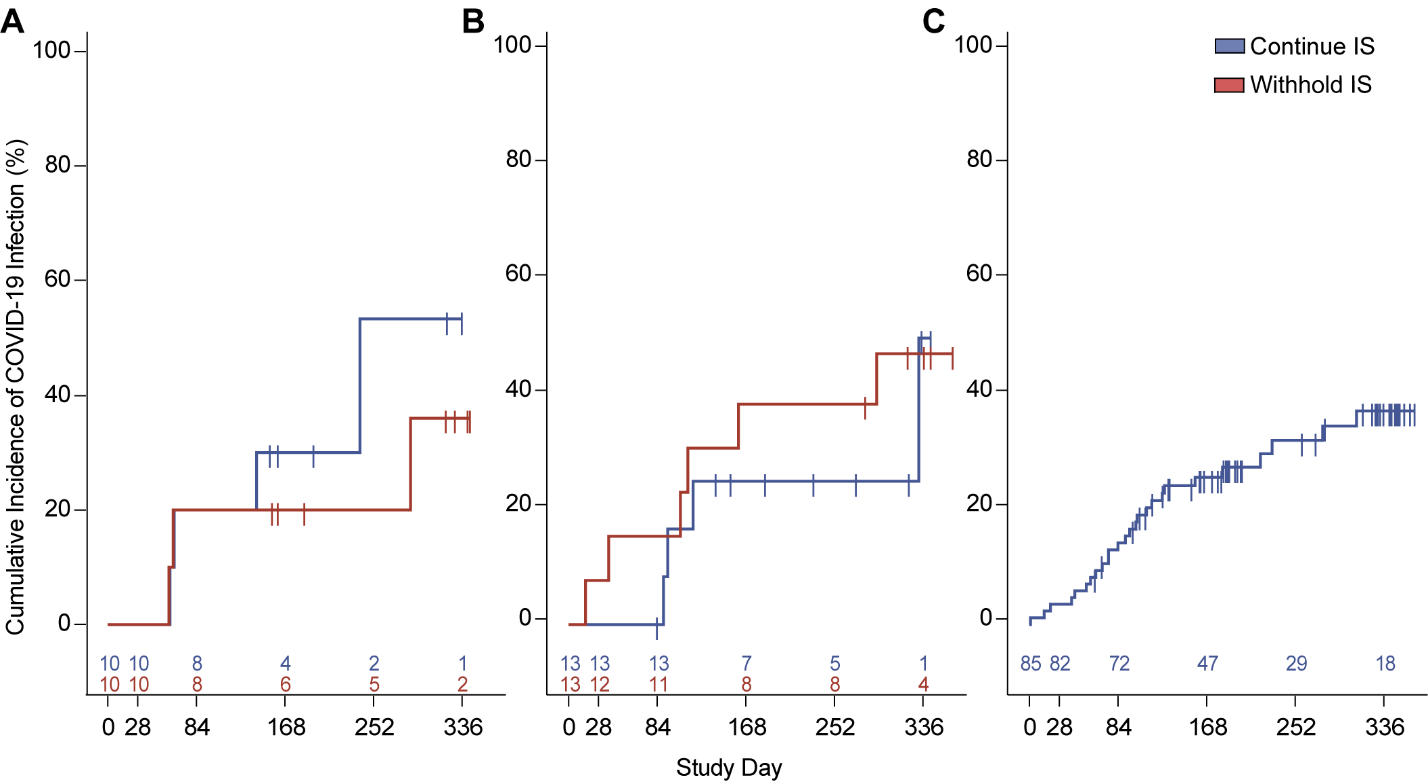
**Supplemental Figure 9. Cumulative incidence of COVID-19 infection in the vaccinated population that received a third mRNA vaccine.** Cumulative incidence of COVID-19 infection in (**A**) mycophenolate mofetil/mycophenolic acid (MMF/MPA)-, (**B**) methotrexate (MTX)-, and (**C**) B cell-depleting therapy (BCDT)-treated autoimmune disease patients throughout 48 weeks post-third vaccination. The cumulative incidence of COVID-19 infections was calculated using the Kaplan-Meier product limit estimator. Vertical lines indicate the timing of censored events.

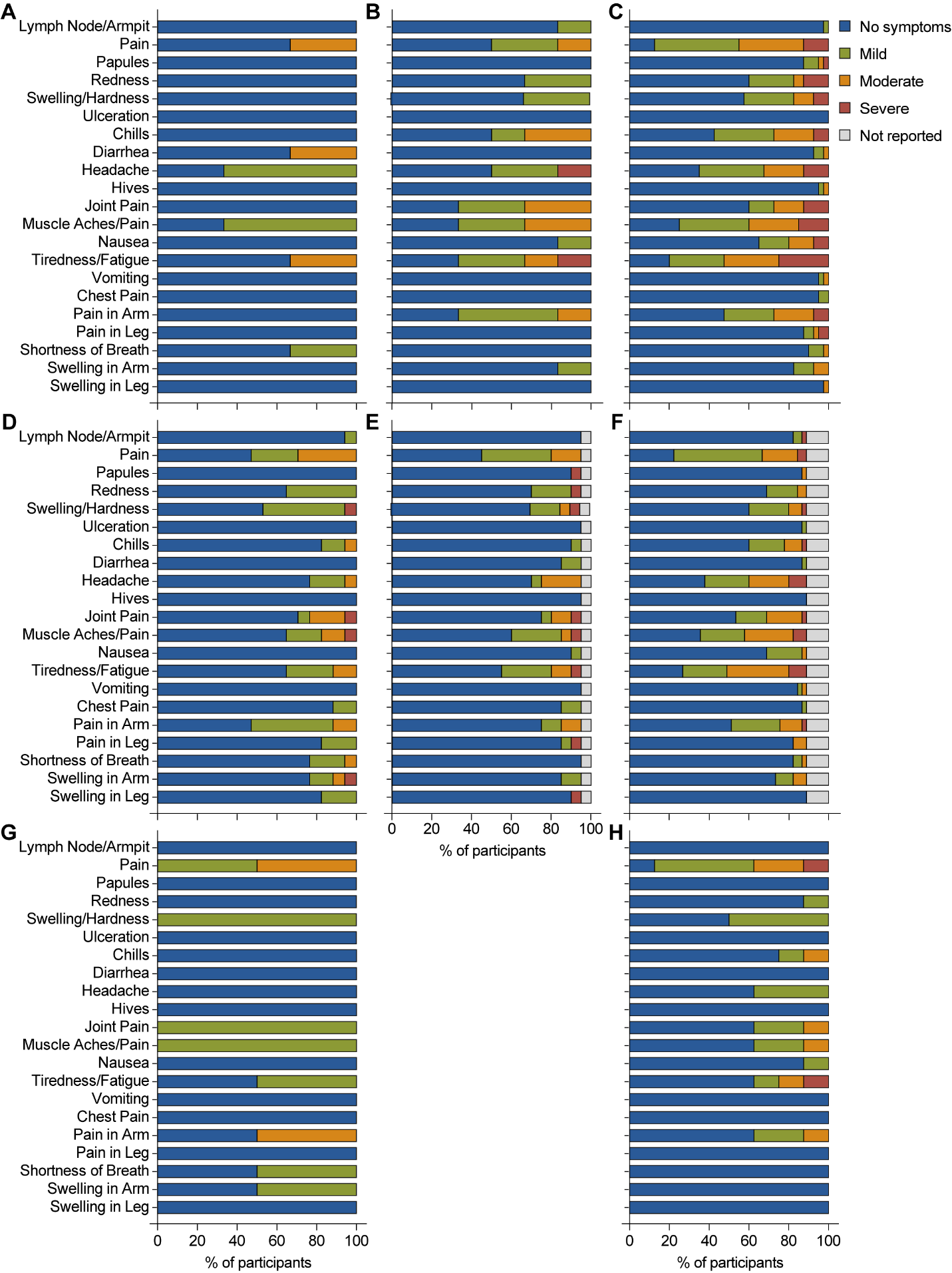
**Supplemental Figure 10. Solicited adverse events following additional vaccination.** Maximum severity of solicited local and systemic reactions over 7 days post-additional (**A-C**) mRNA-1273, (**D-F**) BNT162b2, or (**G-H**) AD26.COV2.S vaccination in (**A, D,** and **G**) mycophenolate mofetil/mycophenolic acid (MMF/MPA)-, (**B** and **E**) methotrexate (MTX)-, and (**C, F,** and **H**) B cell-depleting therapy (BCDT)-treated autoimmune disease patients.

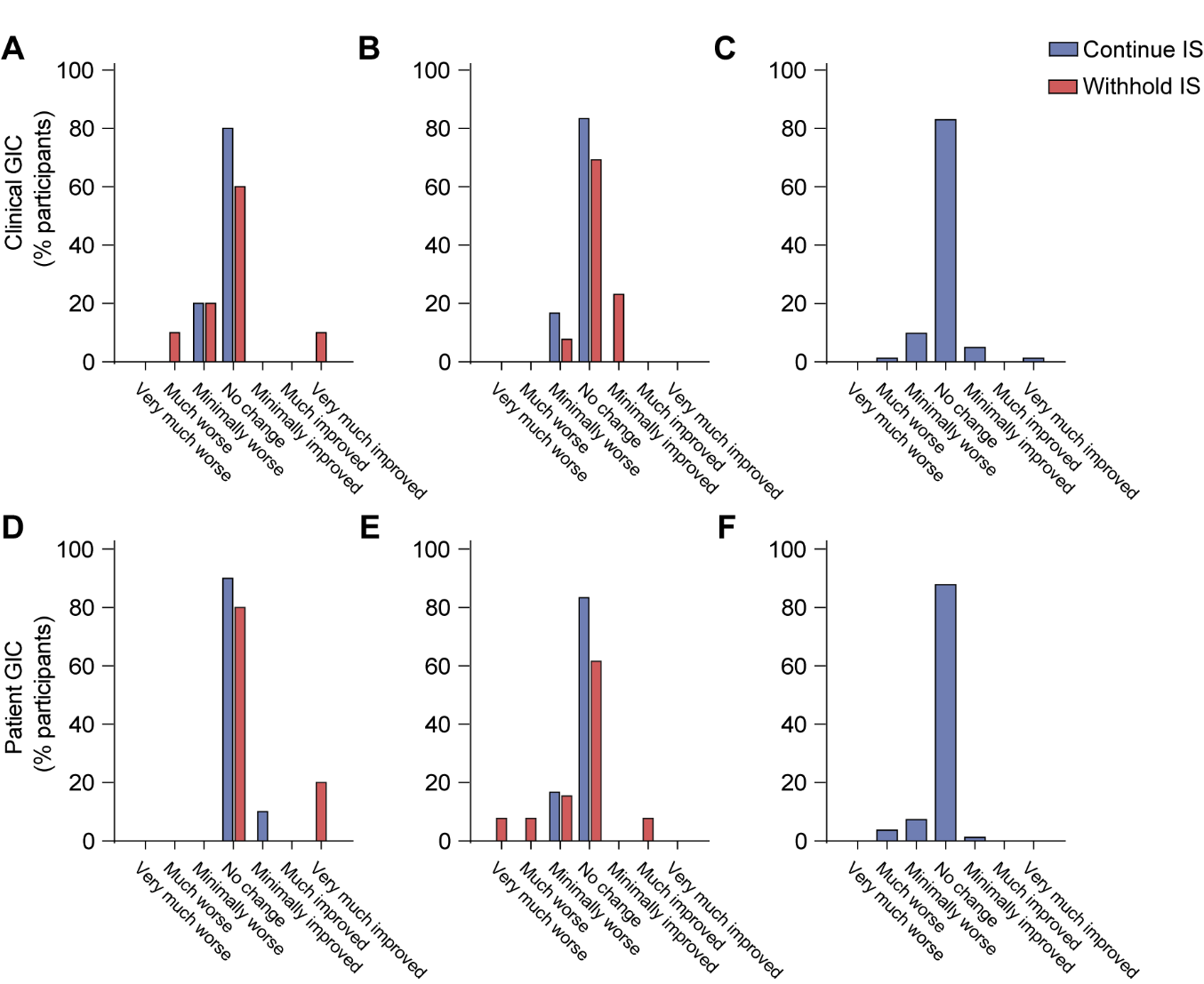

**Supplemental Figure 11. Changes in disease activity in the vaccinated population that received a third mRNA vaccine.** (**A-C**) Clinical and (**D-F**) patient global impression of change (GIC) at week 4 post-third mRNA vaccine in (**A** and **D**) mycophenolate mofetil/mycophenolic acid-, (**B** and **E**) methotrexate-, and (**C** and **F**) B cell-depleting therapy-treated autoimmune disease patients.

**Supplemental Table 1.** Baseline demographics and characteristics by disease in the vaccinated population.

|  | **SLE** | **RA** | **MS** | **SSc** | **Pemphigus** |
| --- | --- | --- | --- | --- | --- |
|  | (n=27) | (n=38) | (n=67) | (n=6) | (n=3) |
| **Sex, n (%)** |  |  |  |  |  |
| Female | 24 (89) | 36 (95) | 44 (66) | 6 (100) | 1 (33) |
| Male | 3 (11) | 2 (5) | 23 (34) | 0 | 2 (67) |
| **Race, n (%)** |  |  |  |  |  |
| White | 13 (48) | 25 (66) | 60 (90) | 5 (83) | 2 (67) |
| Black | 13 (48) | 6 (16) | 6 (9) | 1 (17) | 1 (33) |
| Asian | 0 (0) | 3 (8) | 0 (0) | 0 (0) | 0 (0) |
| Native Hawaiian or Other Pacific Islander | 0 (0) | 0 (0) | 0 (0) | 0 (0) | 0 (0) |
| American Indian or Alaska Native | 0 (0) | 1 (3) | 0 (0) | 0 (0) | 0 (0) |
| Multiple | 0 (0) | 2 (5) | 0 (0) | 0 (0) | 0 (0) |
| Unknown | 1 (4) | 1 (3) | 1 (2) | 0 (0) | 0 (0) |
| **Ethnicity, n (%)** |  |  |  |  |  |
| Hispanic or Latino | 6 (22) | 3 (8) | 5 (8) | 2 (33) | 0 (0) |
| Not Hispanic or Latino | 21 (78) | 35 (92) | 62 (93) | 4 (67) | 3 (100) |
| **Age (years),** mean (SD) | 49.1 (15.4) | 63.0 (11.7) | 45.8 (11.3) | 55.8 (15.6) | 69.7 (6.8) |
| **Response to initial COVID-19 vaccination^a^,** n (%) |  |  |  |  |  |
| Negative | 9 (33) | 17 (45) | 48 (72) | 3 (50) | 2 (67) |
| Sub-optimal | 18 (67) | 21 (55) | 19 (28) | 3 (50) | 1 (33) |
| **Weeks since initial COVID-19 vaccination^b^,** mean (SD) | 23.1 (8.6) | 25.2 (8.1) | 25.5 (5.5) | 29.8 (5.6) | 31.1 (3.1) |
| **Prior COVID-19 infection,** n (%) | 2 (7) | 1 (3) | 4 (6) | 1 (17) | 0 (0) |
| **Vaccine received,** n (%) |  |  |  |  |  |
| BNT162b2 | 21 (78) | 22 (58) | 34 (51) | 4 (67) | 1 (33) |
| mRNA-1273 | 4 (15) | 16 (42) | 26 (39) | 1 (17) | 2 (67) |
| AD26.COV2.S | 2 (7) | 0 (0) | 7 (10) | 1 (17) | 0 (0) |
| **MMF-treated**, n | 16 | 1 | 0 | 3 | 1 |
| mg/day, mean (SD) | 2375 (806.2) | 1000 (-) | NA | 1833 (1040.8) | 3000 (-) |
| **MPA-treated**, n | 2 | 0 | 0 | 2 | 0 |
| mg/day, mean (SD) | 900.0 (763.7) | NA | NA | 2160 (0.0) | NA |
| **MTX-treated**, n | 6 | 25 | 0 | 0 | 0 |
| mg/week, mean (SD) | 21.3 (3.5) | 17.1 (5.6) | NA | NA | NA |
| **Ocrelizumab-treated**, n | 0 | 0 | 62 | 0 | 0 |
| mg/6 months, mean (SD) | NA | NA | 585.5 (64.9) | NA | NA |
| **Ofatumumab-treated**. N | 0 | 0 | 3 | 0 | 0 |
| mg/month, mean (SD) | NA | NA | 20.0 (0.0) | NA | NA |
| **Rituximab-treated**, n | 5 | 18 | 2 | 1 | 2 |
| mg, mean (SD) | 1000 (0.0) | 972.2 (117.9) | 1000 (0.0) | 1000 (-) | 1000 (0.0) |
| **Prednisone-treated**, n | 14 | 6 | 0 | 0 | 1 |
| Mg/day, mean (SD) | 6.0 (3.4) | 4.1 (2.4) | NA | NA | 5.0 (-) |
| **Physician's Global Assessment**, mean (SD) | 1.6 (1.8) | 1.6 (1.5) | 1.5 (2.6) | 1.6 (1.0) | 0.4 (0.6) |
| **Patient's Global Assessment**, mean (SD) | 4.4 (2.9) | 4.2 (2.7) | 3.2 (2.7) | 4.9 (2.1) | 3.5 (3.2) |
| **SLEDAI**, mean (SD) | 3.3 (4.0) | NA | NA | NA | NA |
| **DAS28-CRP**, mean (SD) | NA | 2.9 (1.3) | NA | NA | NA |
| **EDSS**, mean (SD) | NA | NA | 3.0 (2.0) | NA | NA |
| **mRSS**, mean (SD) | NA | NA | NA | 6.2 (10.8) | NA |
| **PDAI**, mean (SD) | NA | NA | NA | NA | 0.3 (0.6) |
| ^a^Negative defined as a Roche Elecsys® Anti-SARS-CoV-2 S result ≤0.79 U/mL and sub-optimal defined as a Roche Elecsys® Anti-SARS-CoV-2 S result >0.79 and ≤ 200 U/mL following the initial COVID-19 vaccine regimen  ^b^Defined as weeks since completed initial course of vaccine to informed consent  DAS28-CRP, Disease Activity Score-28 for Rheumatoid Arthritis with CRP; EDSS, Kurtzke Expanded Disability Status Scale; MMF, mycophenolate mofetil; MPA, mycophenolic acid; mRSS, modified Rodnan Skin Score; MS, multiple sclerosis; MTX, methotrexate; PDAI, Pemphigus Disease Area Index; RA, rheumatoid arthritis; SLE, systemic lupus erythematosus; SLEDAI, Systemic Lupus Erythematosus Disease Activity Index; SSc, systemic sclerosis | | | | | |

**Supplemental Table 2.** Baseline associations with week 4 anti-RBD response in the vaccinated population that received an mRNA vaccine.

|  | **MMF/MPA or MTX**  (n=43) | | **BCDT**  (n=80) | |
| --- | --- | --- | --- | --- |
|  | GM (CV) | p-value^#^ | GM (CV) | p-value^a^ |
| **Vaccine Type** |  |  |  |  |
| mRNA-1273 | 1,510 (515.4) | 0.46 | 9 (943.1) | 0.37 |
| BNT162b2 | 824 (1,312.3) |  | 8 (3,104.6) |  |
| **Immunosuppressant treatment** |  | 0.12 |  |  |
| Withhold | 1,352 (1,302.0) |  | NA |  |
| Continue | 635 (835.4) |  | NA |  |
| **Sex** |  | 0.94 |  | 0.8 |
| Male | 1,975 (45.1) |  | 7 (1,543.9) |  |
| Female | 884 (1,247.1) |  | 9 (1,952.7) |  |
| **Age** |  | 0.61 |  | 0.76 |
| ≤ Median (51 years) | 1,253 (967.0) |  | 7 (1,861.2) |  |
| > Median (51 years) | 812 (1,178.3) |  | 10 (1,732.1) |  |
| **Disease Type** |  | **0.026** |  | 0.47 |
| Systemic Lupus Erythematosus | 788 (2,068.6) |  | 4 (6,828.3) |  |
| Rheumatoid Arthritis | 2,483 (184.5) |  | 4 (683.2) |  |
| Multiple Sclerosis | - |  | 10 (1,951.0) |  |
| Systemic Sclerosis | 73 (148.8) |  | 49 (-) |  |
| Pemphigus | 57 (-) |  | 21 (-) |  |
| **Weeks since original COVID-19 vaccination** |  | 0.55 |  | 0.44 |
| ≤ Median (25.1 weeks) | 832 (829.5) |  | 7 (1,033.0) |  |
| > Median (25.1 weeks) | 1,017 (1,395.7) |  | 11 (3,320.5) |  |
| **Prior COVID-19 Infection?** |  | 0.24 |  | 0.36 |
| Yes | 6,504 |  | 17 (1,851.5) |  |
| No | 893 (1,079.0) |  | 8 (1,783.2) |  |
| **Prednisone use** |  | 0.67 |  | 0.77 |
| Yes | 1,585 (240.8) |  | 15 (12,149.3) |  |
| No | 725 (2,025.7) |  | 8 (1,530.1) |  |
| **Medication** |  | 0.17 |  | 0.23 |
| MMF/MPA | 520 (2,494.9) |  | NA |  |
| MTX | 1,556 (417.8) |  | NA |  |
| Rituximab | NA |  | 5 (1,289.3) |  |
| Ocrelizumab | NA |  | 10 (1,974.2) |  |
| Ofatumumab | NA |  | 23 (3,390.9) |  |
| **Baseline CD19 count** |  |  |  | **0.0014** |
| Absent | NA |  | 4 (741.1) |  |
| Present | NA |  | 29 (3,204.9) |  |
| **Days since last BCDT dose** |  |  |  | 0.13 |
| ≤ Median (127 days) | NA |  | 5 (536.4) |  |
| > Median (127 days) | NA |  | 14 (4,566.9) |  |
| BCDT, B cell-depleting therapy; CV, coefficient of variation; GM, geometric mean; MMF/MPA, mycophenolate mofetil/mycophenolic acid; MTX, methotrexate  ^a^Wilcoxon rank-sum test, bold text denotes statistical significance (p<0.05) | | | | |

**Supplemental Table 3.** Incidence of adverse events (AEs) in participants who received a third mRNA vaccine.

|  | **MMF/MPA** | | | | **MTX** | | | | **BCDT** | |
| --- | --- | --- | --- | --- | --- | --- | --- | --- | --- | --- |
|  | **Continue** | | **Withhold** | | **Continue** | | **Withhold** | |  |  |
|  | Subjects (n=10) | Events | Subjects (n=10) | Events | Subjects (n=13) | Events | Subjects (n=13) | Events | Subjects (n=85) | Events |
| **AEs** | 4 (40) | 6 | 4 (40) | 5 | 5 (39) | 7 | 6 (46) | 6 | 39 (46) | 55 |
| Grade 1 | 4 (40) | 5 (83) | 3 (30) | 3 (60) | 1 (8) | 1 (14) | 2 (15) | 2 (33) | 19 (22) | 20 (36) |
| Grade 2 | 1 (10) | 1 (17) | 1 (10) | 2 (40) | 4 (31) | 6 (86) | 3 (23) | 3 (50) | 20 (24) | 22 (40) |
| Grade 3 | 0 (0) | 0 (0) | 0 (0) | 0 (0) | 0 (0) | 0 (0) | 1 (8) | 1 (17) | 10 (12) | 13 (24) |
| Grade 4 | 0 (0) | 0 (0) | 0 (0) | 0 (0) | 0 (0) | 0 (0) | 0 (0) | 0 (0) | 0 (0) | 0 (0) |
| Grade 5 | 0 (0) | 0 (0) | 0 (0) | 0 (0) | 0 (0) | 0 (0) | 0 (0) | 0 (0) | 0 (0) | 0 (0) |
| Related to Vaccine | 0 (0) | 0 (0) | 0 (0) | 0 (0) | 0 (0) | 0 (0) | 0 (0) | 0 (0) | 6 (7) | 6 (11) |
| **SAEs** | 0 (0) | 0 | 0 (0) | 0 | 0 (0) | 0 | 1 (8) | 1 | 9 (11) | 11 |
| Related to Vaccine | 0 (0) | 0 (0) | 0 (0) | 0 (0) | 0 (0) | 0 (0) | 0 (0) | 0 (0) | 1 (1) | 1 (9) |
| **MAAE^a^** | 0 (0) | 0 (0) | 0 (0) | 0 (0) | 0 (0) | 0 (0) | 0 (0) | 0 (0) | 1 (1) | 1 (2) |
| **NOCMC^b^** | 0 (0) | 0 (0) | 0 (0) | 0 (0) | 0 (0) | 0 (0) | 0 (0) | 0 (0) | 7 (8) | 7 (13) |
| **AESI** |  |  |  |  |  |  |  |  |  |  |
| Myocarditis | 0 (0) | 0 (0) | 0 (0) | 0 (0) | 0 (0) | 0 (0) | 0 (0) | 0 (0) | 0 (0) | 0 (0) |
| Pericarditis | 0 (0) | 0 (0) | 0 (0) | 0 (0) | 0 (0) | 0 (0) | 0 (0) | 0 (0) | 0 (0) | 0 (0) |

Values depict n (%)

Percentages for the number of participants with events are based on the number of participants in the vaccinated population.
AE, adverse event; AESI, adverse events of special interest; MAAE, medically attended adverse event; NOCMC, new-onset chronic medical condition; SAE, serious adverse event
^a^MAAE defined as a hospitalization, emergency room visit, or unscheduled medical visit for any reason and considered related to the vaccine. ^b^NOCMC is defined as any new ICD diagnosis during the study after receipt of the vaccine that will continue for at least 3 months and requires continued health care intervention.

**Supplemental Table 4.** Incidence of adverse events (AEs) in participants who received a second AD26.COV2.S vaccine.

|  | **MMF/MPA** | | **BCDT** | |
| --- | --- | --- | --- | --- |
|  | **Withhold** | |  |  |
|  | Subjects (n=2) | Events | Subjects (n=8) | Events |
| **AEs** | 1 (50) | 1 | 6 (75) | 6 |
| Grade 1 | 0 (0) | 0 (0) | 4 (50) | 4 (67) |
| Grade 2 | 0 (0) | 0 (0) | 2 (25) | 2 (33) |
| Grade 3 | 1 (100) | 1 (100) | 0 (0) | 0 (0) |
| Grade 4 | 0 (0) | 0 (0) | 0 (0) | 0 (0) |
| Grade 5 | 0 (0) | 0 (0) | 0 (0) | 0 (0) |
| Related to Vaccine | 0 (0) | 0 (0) | 0 (0) | 0 (0) |
| **SAEs** | 1 (50) | 1 | 0 (0) | 0 |
| Related to Vaccine | 0 (0) | 0 (0) | 0 (0) | 0 (0) |
| **MAAE^a^** | 0 (0) | 0 (0) | 0 (0) | 0 (0) |
| **NOCMC^b^** | 0 (0) | 0 (0) | 0 (0) | 0 (0) |
| **AESI** |  |  |  |  |
| Myocarditis | 0 (0) | 0 (0) | 0 (0) | 0 (0) |
| Pericarditis | 0 (0) | 0 (0) | 0 (0) | 0 (0) |

Values depict n (%)

Percentages for the number of participants with events are based on the number of participants in the vaccinated population.
AE, adverse event; AESI, adverse events of special interest; MAAE, medically attended adverse event; NOCMC, new-onset chronic medical condition; SAE, serious adverse event; VAE, vaccine-related adverse event
^a^MAAE defined as a hospitalization, emergency room visit, or unscheduled medical visit for any reason and considered related to the vaccine.
^b^NOCMC is defined as any new ICD diagnosis during the study after receipt of the vaccine that will continue for at least 3 months and requires continued health care intervention.

**Supplemental** **Table 5.** Disease activity at 4 weeks post-booster vaccination in participants who received a third mRNA vaccine.

|  | **MMF/MPA** | | **MTX** | | **BCDT** | **All** |
| --- | --- | --- | --- | --- | --- | --- |
|  | Continue | Withhold | Continue | Withhold |  |  |
| **SLE participants**, n | 7 | 8 | 3 | 3 | 4 | 25 |
| Severe flare^a^, n (%) | 0 (0) | 0 (0) | 0 (0) | 0 (0) | 0 (0) | 0 (0) |
| Mild/moderate flare^1^, n (%) | 3 (43) | 1 (13) | 1 (33) | 0 (0) | 0 (0) | 5 (20) |
| **RA participants**, n | 0 | 0 | 9 | 10 | 17 | 36 |
| Severe flare^b^, n (%) | NA | NA | 0 (0) | 0 (0) | 1 (6) | 1 (3) |
| Moderate flare^2^, n (%) | NA | NA | 1 (11) | 3 (30) | 8 (47) | 12 (33) |
| **MS participants**, n | 0 | 0 | 0 | 0 | 60 | 60 |
| Relapse^c^, n (%) | NA | NA | NA | NA | 0 (0) | 0 (0) |
| **SSc participants**, n | 3 | 1 | 0 | 0 | 1 | 5 |
| Severe flare^d^, n (%) | 0 (0) | 0 (0) | NA | NA | 0 (0) | 0 (0) |
| **Pemphigus participants**, n | 0 | 1 | 0 | 0 | 2 | 3 |
| Severe flare^e^, n (%) | NA | 1 (100) | NA | NA | 0 (0) | 1 (33) |
| **All participants,** n | 10 | 10 | 12 | 13 | 84 | 129 |
| Severe flare, n (%) | 0 (0) | 1 (10) | 0 (0) | 0 (0) | 1 (1) | 2 (2) |
| BCDT, B cell-depleting therapy; MMF/MPA, mycophenolate mofetil/mycophenolic acid; MTX, methotrexate; MS, multiple sclerosis; RA, rheumatoid arthritis; SSc, systemic sclerosis; SLE, systemic lupus erythematosus | | | | | | |
| Disease activity measured by ^a^SELENA-SLEDAI; ^b^DAS-28 CRP; ^c^Kurtzke Expanded Disability Status Scale; ^d^modified Rodnan Skin Score; ^d^Pemphigus Disease Area Index | | | | | | |

**Supplemental Table 6.** Participating study centers

| **Site Name** | **Site PI** | **Screened** | **Randomized/ Allocated** |
| --- | --- | --- | --- |
| Benaroya Research Institute | Sandra Lord, MD | 4 | 4 |
| Brigham and Women’s Hospital | Jeffrey A. Sparks, MD, MMSc | 12 | 8 |
| Cleveland Clinic | Jeffrey Cohen, MD | 19 | 11 |
| Columbia University | Anca Askanase, MD | 10 | 0 |
| Duke University Medical Center | Ankoor Shah, MD | 1 | 0 |
| Emory University | Arezou Khosroshahi, MD | 42 | 10 |
| Feinstein Institute for Medical Research | Meggan Mackay, MD | 42 | 34 |
| Massachusetts General Hospital – Division of Rheumatology | Zachary Wallace, MD | 18 | 11 |
| Michigan Medicine | Dinesh Khanna, MD | 29 | 19 |
| Medical University of South Carolina | Diane L. Kamen, MD | 9 | 1 |
| New York University Langone Orthopedic Center | Amit Saxena, MD | 10 | 4 |
| Oklahoma Medical Research Foundation | Judith James, MD, PhD | 47 | 25 |
| Temple University, Lewis Katz SOM | Roberto Caricchio, MD | 5 | 4 |
| University of California, Los Angeles Division of Rheumatology | Maureen McMahon, MD | 0 | 0 |
| University of Pennsylvania Perelman Center for Advanced Medicine | Amit Bar-Or, MD | 26 | 14 |
| University of Rochester Medical Center | Christopher Palma, MD, ScM | 0 | 0 |
| University of Texas Health Science Center at Houston | Maureen D. Mayes, MD | 0 | 0 |
| Washington University School of Medicine | Alfred Kim, MD, PhD | 3 | 2 |
| Yale University School of Medicine | Fotios Koumpouras, MD | 2 | 1 |

**Supplemental Acknowledgements**

| **ACV01 Study Team** | **Institution** |
| --- | --- |
| Alise Carlson, MD | Cleveland Clinic |
| Deja Rose, MD | Cleveland Clinic |
| Lindsay Ross, MD | Cleveland Clinic |
| Sarah Simmons, MD | Cleveland Clinic |
| Ahmad Mahadeen, MD | Cleveland Clinic |
| Megan Elder, RN BSN | Cleveland Clinic |
| Leah Tardivo, RN BSN | Cleveland Clinic |
| Bryan Davies, RN | Cleveland Clinic |
| Diane Ivancic | Cleveland Clinic |
| Stephanie Moore | Cleveland Clinic |
| Sadie Coates | Cleveland Clinic |
| Michelle Maxson | Cleveland Clinic |
| Rachael Yim | Cleveland Clinic |
| Jenna Titus | Cleveland Clinic |
| Ariel S. Bizon | Cleveland Clinic |
| Alecia Chase, RN BSN | Cleveland Clinic |
| Moein Amin, MD | Cleveland Clinic |
| Lisa Stropp, MD | Cleveland Clinic |
| Jameson Holloman, MD | Cleveland Clinic |
| Megan Roser | Cleveland Clinic |
| Jacqueline Dinishak | Cleveland Clinic |
| Amanda Evans | Cleveland Clinic |
| Thomai Skaramagas | Cleveland Clinic |
| Cristina Arriens, MD | Oklahoma Medical Research Foundation |
| Teresa Aberle, PA-C | Oklahoma Medical Research Foundation |
| Kelli Kraus | Oklahoma Medical Research Foundation |
| Micki Drake | Oklahoma Medical Research Foundation |
| Jason L. Larabee | University of Oklahoma Health Sciences |
| Patricia Gonzales, LVN | University of Texas Health Science Center |
| Julio Charles, MS | University of Texas Health Science Center |
| Asaff Harel, MD | Northwell Health |
| Erik Anderson, MD, PhD | Northwell Health |
| Cynthia Aranow, MD | Northwell Health |
| Janine Sullivan, NP | NYU Langone Health |
| Monica Gamez-Perez, RN | NYU Langone Health |
| Emilie Schurenberg | NYU Langone Health |
| Abraham Sinay-Smith | NYU Langone Health |
